# Time-specific bidirectional links between maternal microbiome, milk composition and infant gut microbiota

**DOI:** 10.64898/2025.12.11.25342041

**Authors:** Lishi Deng, Kelsey Fehr, Laeticia Celine Toe, Lindsay H. Allen, Lars Bode, Daniela Hampel, Melissa B. Manus, Andrew Mertens, Bianca Robertson, Chloe Yonemitsu, Bruno De Meulenaer, Carl Lachat, Justin L. Sonnenburg, Meghan B. Azad, Trenton Dailey-Chwalibóg

**Affiliations:** Department of Food Technology, Safety and Health, Faculty of Bioscience Engineering, Ghent University, Ghent, Belgium; Department of Pediatrics and Child Health, University of Manitoba, Winnipeg, Canada; Manitoba Interdisciplinary Lactation Centre (MILC), Children’s Hospital Research Institute of Manitoba, Winnipeg, Canada; Institut de Recherche en Sciences de la Santé (IRSS), Unité Nutrition et Maladies Métaboliques; USDA, ARS Western Human Nutrition Research Center, University of California, Davis, CA; Department of Pediatrics,and Larsson-Rosenquist Foundation Mother-Milk-Infant Center of Research Excellence (MOMI CORE), and Human Milk Institute (HMI), University of California San Diego, La Jolla, CA, USA; Institute for Global Nutrition, Department of Nutrition, University of California, Davis, CA; Department of Anthropology, University of Texas, San Antonio, USA; School of Public Health University of California, Berkeley, USA; Department of Microbiology and Immunology, Stanford University School of Medicine, Stanford, CA, USA; Chan Zuckerberg Biohub, San Francisco, CA, USA; Center for Human Microbiome Studies, Stanford University School of Medicine, Stanford, CA, USA; Agence de Formation de Recherche et d’Expertise en Santé pour l’Afrique (AFRICSanté), Bobo-Dioulasso, Burkina Faso

## Abstract

Early-life gut microbiome development is shaped by complex maternal and nutritional influences, yet the temporal and directional structure of these interactions remains unclear. In a longitudinal study of 152 mother-infant dyads in rural Burkina Faso, we examined how maternal gut and milk microbiomes, alongside milk components, influence infant gut microbiome development during the first six months. At 1–2 months, the infant gut microbiome clustered into three types: *Escherichia*-dominated, *Bifidobacterium*-dominated, and a diverse, pathogen-prevalent profile, which became less distinct by 5–6 months. Early infant gut microbiomes were associated with maternal prenatal gut microbiota and early milk microbiome and oligosaccharides, while later variation was linked to other milk nutrients. Furthermore, early infant gut profiles predicted subsequent milk composition, suggesting potential bidirectional communication between infant needs and maternal lactational physiology. These findings offer new insights into early-life microbial development and inform future mechanistic studies and microbiome-targeted interventions, particularly in low-resource settings.

## Introduction

The colonization and evolution of the infant gut microbiome inform immune development, metabolic programming, and long-term health outcomes in early life and through adulthood^1–3^. The dynamic microbial colonization of the gut is driven by a complex interplay of maternal, environmental and nutritional factors^4–6^. However, the timing, coordination and relative contributions of these factors, and how they interact to shape microbial trajectories across infancy, remain incompletely understood. This question is particularly relevant for low- and middle-income countries (LMICs), where rapid shifts in the gut microbiome during early life often occur alongside environmental exposure (e.g., to pathogens or poor sanitation), infectious disease burden, and nutritional vulnerabilities^7–9^. Identifying time-specific drivers of microbial succession is therefore critical to inform strategies for optimizing infant health.

The maternal microbiome serves as a key source of microbial seeding during the first weeks to months of life^10^. Vertical transmission of vaginal and gut microbes occurs during delivery^11^, while maternal skin and oral exposures continue to shape the infant gut through frequent contact in the early months of life^12,13^. In parallel, breastfeeding introduces both commensal bacteria and a broad array of bioactive components that can modulate gut microbial composition by promoting or displacing specific taxa^14–16^. Among these, human milk oligosaccharides (HMOs) have been widely studied as selective substrates for beneficial microbes, including *Akkermansia* and certain *Bifidobacterium* and *Bacteroides* species^15,17–20^.

Yet, human milk is far more than a source of HMOs. It is a complex, dynamic biomatrix containing macronutrients, micronutrients, hormones, and metabolites that vary across lactation and between individuals^21,22^. These components reflect both maternal physiological status and infants’ developmental need^23–25^, and emerging evidence suggests that they may shape microbial colonization patterns in parallel with, or independent of, HMOs^16,26,27^. However, most studies to date have focused narrowly on individual milk components or single time points, leaving major gaps in understanding how shifts in milk composition interact with the developing infant gut microbiome during early life—a critical window for microbiome-mediated immune maturation and growth that underpins infant health and survival.

Moreover, few studies have integrated milk and gut microbiome data in a longitudinal, time-resolved manner across early lactation. Particularly understudied is the potential for bidirectional communication whereby maternal physiology shapes early colonization, while early infant gut patterns may, in turn, modulate milk composition later in lactation via signaling pathways or infant-driven cues^24,28^. For instance, immune activation in the infant may trigger changes in milk immunoglobulin or cytokine levels through maternal-infant feedback mechanisms^29^. Such temporal feedback loops likely play an important role in aligning maternal milk output with infant need, not only in terms of nutrition, but also in response to immune or microbial signals.

To address these gaps, we conducted a longitudinal, multi-omics analysis of milk and maternal and infant stool samples from the Micronutriments pour la SAnté de la Mère et de l’Enfant-III (MISAME-III) randomized controlled trial in rural Burkina Faso. The MISAME-III trial evaluated the efficacy of balanced energy–protein (BEP) supplementation during pregnancy and lactation on birth outcomes and infant growth^30–32^. Within this trial, we profiled the maternal gut microbiome during pregnancy and postpartum, the milk microbiome, a broad panel of milk components (including macronutrients, minerals, vitamins, bioactive compounds, and metabolites), and the infant gut microbiome at multiple time points over the first six months^33^. In earlier work, we found that BEP supplementation altered the maternal gut microbiome and enhanced microbial carbohydrate metabolism in the infant gut^34^.

Here, we characterize early infant gut microbiome clusters and their changes over time, identify maternal and milk-related determinants of these patterns, and explore how early gut microbiome profiles may, in turn, influence milk composition later in lactation.

## Results

### Study cohort, microbial community composition, and longitudinal milk sampling

This study included a subset of 152 mother-infant dyads (Figure 1A). Maternal stool samples (*n* = 472 total) were collected at four time points: second trimester (Tri2, *n* = 42), third trimester (Tri3, *n* = 149), 1–2 months (*n* = 139), and 5–6 months postpartum (*n* = 142). Maternal milk samples (*n* = 423 total) were collected at 14–21 days (*n* = 138), 1–2 months (*n* = 144), and 3–4 months (*n* = 141). Infant stool samples (*n* = 256 total) were collected at 1–2 months (*n* = 142) and 5–6 months (*n* = 114), corresponding to the maternal postnatal time points.

**Figure 1.**
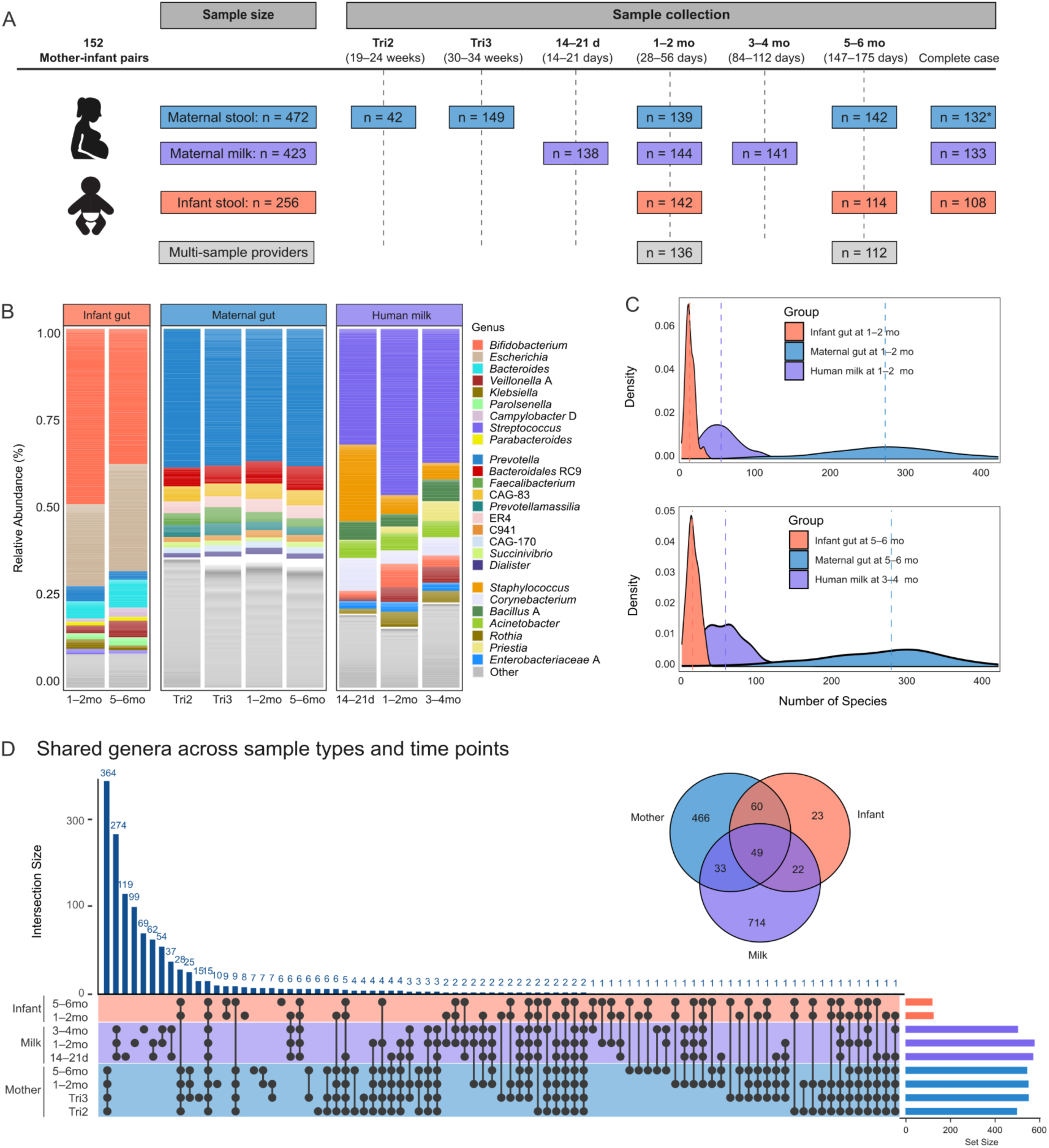
Longitudinal sampling design and microbial community structures across infant and maternal gut, and human milk microbiomes. **(A)** Sampling timeline and sample size for maternal stool, breast milk, and infant stool collections across the study period. Maternal stool samples from the second trimester (Tri2) were excluded from complete-case calculation due to limited sampling. **(B)** Relative abundances (%) of the top ten microbial genera in infant gut, maternal gut, and breast milk microbiomes, aggregated by sample collection time point. Genera outside the top ten are grouped under “Other.” **(C)** Species richness (observed species count) across three compartments at 1–2 months (top panel), and at later time points (3–4 months for milk and 5–6 months for infant and maternal gut; bottom panel). **(D)** Shared and unique microbial genera across sample types. Upset plot shows intersections between maternal gut, breastmilk and infant gut genera across time points; horizontal bars represent total genera per compartment, vertical bars represent shared genera. Venn diagram summarizes overlaps across the three microbiomes.

Participating mothers had a mean age of 24.3 years (SD ±5.6) and an average body mass index (BMI) of 22.4 (±3.5) kg/m^2^. Infants were born at a mean gestational age of 39.8 (±1.7) weeks, with an average birth weight of 3.0 (±0.47) kg and length of 48.5 (±2.16) cm. Nearly all infants were exclusively breastfed, with an average exclusive breastfeeding duration of 5.78 months (Table S1).

Microbial composition was characterized using deep shotgun metagenomic sequencing for maternal and infant stool samples, and 16S rRNA gene sequencing for milk samples. The gut and milk microbiomes exhibited distinct taxonomic profiles across sample types (Figure 1B). The infant gut microbiome was dominated by *Bifidobacterium* species (median relative abundance 44.7%; prevalence 98.0%), followed by *Escherichia* species (median 15.4%; prevalence 94.0%), with a decline in *Bifidobacterium* and an increase in *Escherichia* observed over time. In the maternal gut, *Prevotella* species were predominant, representing a median of 37.9% of the community, with little variation between pre- and postnatal samples. The maternal milk microbiome was dominated by *Streptococcus* species (median 42.8%, detected in all milk samples), with notable decreases in *Staphylococcus* abundance observed early in lactation.

Microbial richness, based on the number of observed species, varied markedly across sample types (Figure 1C). The infant gut microbiome had notably lower richness (range 2–43 species per sample) compared to maternal milk (8–195 species per sample) and maternal gut samples (75–614 species per sample). Across all samples, we detected a total of 1,367 unique microbial genera in at least one sample. Among these, 714 genera were unique to maternal milk, 466 were unique to the maternal stool samples, and only 23 were exclusive to infant gut (Figure 1D). This suggests that the milk microbiome, despite lower within-sample richness, displayed substantial inter-individual variability. A total of 109 genera were shared between maternal gut and infant gut, 82 between maternal gut and milk, and 71 between maternal milk and infant gut, while only 49 genera were common to all three compartments.

Among the 144 mothers who provided milk samples, 112 were classified as secretors and 32 as non-secretors. Secretor status reflects the presence or absence of fucosyltransferase 2 gene activity, which determines the ability to produce fucosylated HMOs such as 2’-fucosyllactose (2’FL)^35^. As expected, HMO concentrations peaked at 14–21 days postpartum, with total levels significantly higher in secretors (Figure S1).

Macronutrient levels remained relatively stable throughout lactation, except for protein, which declined from an average of 1.49% at 14–21 days to 0.94% at 3–4 months (Figure S2). Several bioactive proteins showed temporal shifts: insulin and luteinizing hormone (LH) increased, while fibroblast growth factor-21 (FGF-21), follicle-stimulating hormone (FSH) and calprotectin decreased over time. Most mineral and vitamin concentrations also declined during lactation. Notably, zinc levels dropped by approximately half from early to late lactation. Other declining components included sodium, phosphorus, potassium, iron, selenium, alpha-tocopherol, vitamin A, vitamin B12, and vitamin B3-related compounds including nicotinamide adenine dinucleotide (NAD), nicotinamide mononucleotide (NMN), nicotinamide (NAM). In contrast, concentrations of magnesium, free thiamin, pyridoxal, and vitamin B6 increased over time.

### Three distinct infant gut microbiome clusters at 1–2 months and their transitions by 5–6 months

To explore variation in early-life microbiome development, we performed hierarchical clustering of infant gut microbiomes at 1–2 months. This revealed three distinct community clusters (Figure 2A), primarily characterized by the dominance of *Escherichia* (cluster 1, C1; *n* = 44), *Bifidobacterium* (cluster2, C2; *n* = 65), and a more compositionally diverse community (cluster 3, C3; *n* = 30). Infants in C3 exhibited higher prevalence of enteric pathogens as measured by quantitative PCR using TaqMan Array Card (TAC), including *Salmonella* (detected via *Salmonella* invasion gene A and tetrathionate reductase gene) and *Adenovirus* (serotypes 40 and 41), compared to those in C1 and C2 (Figure 2B). These findings reflect molecular detection of pathogen DNA, which may represent asymptomatic carriage rather than active clinical infection.

**Figure 2.**
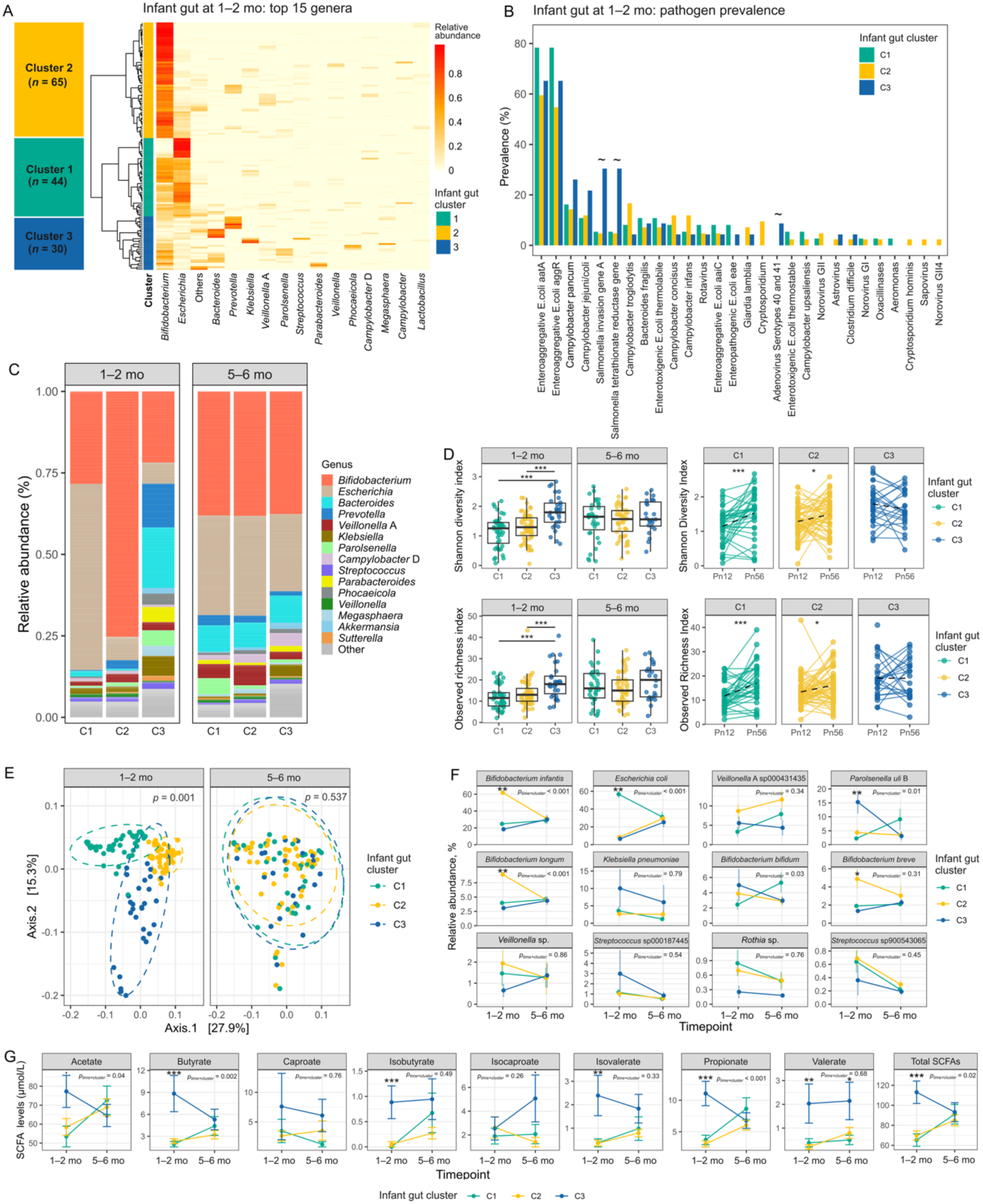
Three distinct infant gut clusters at 1–2 months and their transition by 5–6 months. **(A)** Hierarchical clustering (Bray-Curtis, Ward’s method) heatmap of the top 15 genera in infant stool samples at 1–2 months, identifying three distinct microbial community types. Low-abundance genera were grouped as “Other”. **(B)** Prevalence (%) of enteric pathogens detected by TaqMan Array Card (TAC) in infant stool at 1–2 months. “∼” indicates unadjusted *p* < 0.05 based on Fisher’s exact test before multiple testing correction (Benjamini–Hochberg). **(C)** Stacked-bar plots showing relative abundance (%) of the 15 most dominant genera by cluster at 1–2 and 5–6 months. Genera beyond the top 15 are grouped under “Other.” **(D)** Alpha diversity metrics across clusters and time points. Left panels: Boxplots of Shannon diversity and observed richness, with statistical testing by linear mixed-effects models (LME) including participant as a random effect, followed by pairwise estimated marginal means contrasts. Right panels: Paired within-individual changes from 1–2 to 5–6 months. **(E)** Principal Coordinates Analysis (PCoA) of beta diversity based on Weighted UniFrac distances, showing separation by clusters at 1–2 months. PERMANOVA p-values adjusted using Benjamini–Hochberg correction. **(F)** Temporal trends in relative abundances (%) for the 12 most abundant microbial species across clusters from 1–2 to 5–6 months. Data points represent means with error bars indicating standard error (SE). Differences between clusters within each time point were assessed using ANOVA; interaction p-values (time × clusters) derived from LME models and adjusted for Benjamini–Hochberg correction. **(G)** Temporal changes in short-chain fatty acid (SCFA) concentrations (µmol/L) among the three infant gut clusters defined at 1–2 months. Differences between clusters within each time point were assessed using ANOVA; interaction p-values (time × clusters) derived from LME models and adjusted for Benjamini–Hochberg correction. \*\*\**p_(FDR)_* < 0.001; \*\**p_(FDR)_* < 0.01; \**p_(FDR)_* < 0.05; *·p_(FDR)_* < 0.1.

Maternal and infant baseline characteristics were broadly comparable across infant gut clusters, with two notable exceptions (Figure S3; Table S1). Mothers of C2 infants had higher gestational weight gain compared to those in C3 (7.03 kg vs 4.96 kg, *p_unadjusted_* = 0.04), and households of C2 infants had a higher average number of children under five years old compared to those of C1 (1.33 vs 0.82, *p_unadjusted_* = 0.02). However, these differences did not remain significant after correction for multiple hypothesis testing.

By 5–6 months, infants no longer clustered according to their 1–2-month microbiome types (Figure S4). Most infants (*n* = 88) exhibited a gut profile dominated by *Bifidobacterium* (with or without high *Escherichia* abundance), while a smaller number (*n* = 17) retained a profile dominated by *Escherichia* alone, and a few (*n* = 7) were dominated by *Bacteroides*. The overall infant gut microbiome transitioned toward a composition characterized by mixed dominance of *Bifidobacterium* (∼40%) and *Escherichia* (∼30%), although inter-individual variability remained high (Figure 2C).

Differential abundance analyses supported these changes: species-level distinctions present at 1–2 months, such as enrichment of *Escherichia coli* in C1 and *Bifidobacterium infantis* in C2, were no longer observed by 5–6 months (Figure S5A). Similarly, numerous gene-level differences observed at 1–2 months were absent by 5–6 months (Figure S5B).

Microbial diversity increased over time, particularly among C1 and C2 infants. At 1–2 months, C3 had the highest Shannon diversity and richness, but by 5–6 months, diversity metrics were comparable across all clusters (Figure 2D). Principal Coordinate Analysis (PCoA) supported this pattern, showing clear separation of clusters at 1–2 months (*p_FDR_* = 0.001), but substantial overlap by 5–6 months (*p_FDR_* = 0.537, Figure 2E). The 12 most abundant species showed significant differences between clusters at 1–2 months, driven largely by species such as *B. infants*, *B. longum*, *B. breve*, *E. coli*, and *Parolsenella uli* B (Figure 2F). However, these differences largely diminished at 5–6 months.

Short-chain fatty acid (SCFA) levels also varied by clusters at 1–2 months, with consistently higher levels in C3 infants. By 5–6 months, most differences in SCFAs resolved, except for valerate, which remained elevated in C3 infants (Figure 2G). At 1–2 months, correlation analysis revealed negative associations between several SCFAs and *B. infantis*, *B. longum*, *E. coli*, and *B. fragilis* A, whereas a *Prevotella* species showed positive correlations, particularly with butyrate and valerate (Figure S6). By 5–6 months, negative associations weakened substantially, and *B. bifidum* exhibited positive correlations with multiple SCFAs, potentially suggesting a shift in microbial metabolic activity over time.

### Maternal gut microbiome in the third trimester associated with early infant gut clusters

Subtle but notable variations in the prenatal maternal gut microbiome were associated with infant gut microbiome clusters defined at 1–2 months, primarily evident in the third trimester (Tri3). Mothers of C2 infants had higher relative abundances of *Prevotella* species in their gut microbiome, both during the second (Tri2; 45.7%) and Tri3 (42.6%), compared to lower abundances in C1 (33.5% at Tri2 and 36.8% at Tri3) and C3 (30.4% at Tri2 and 29.3% at Tri3) (Figure 3A).

**Figure 3.**
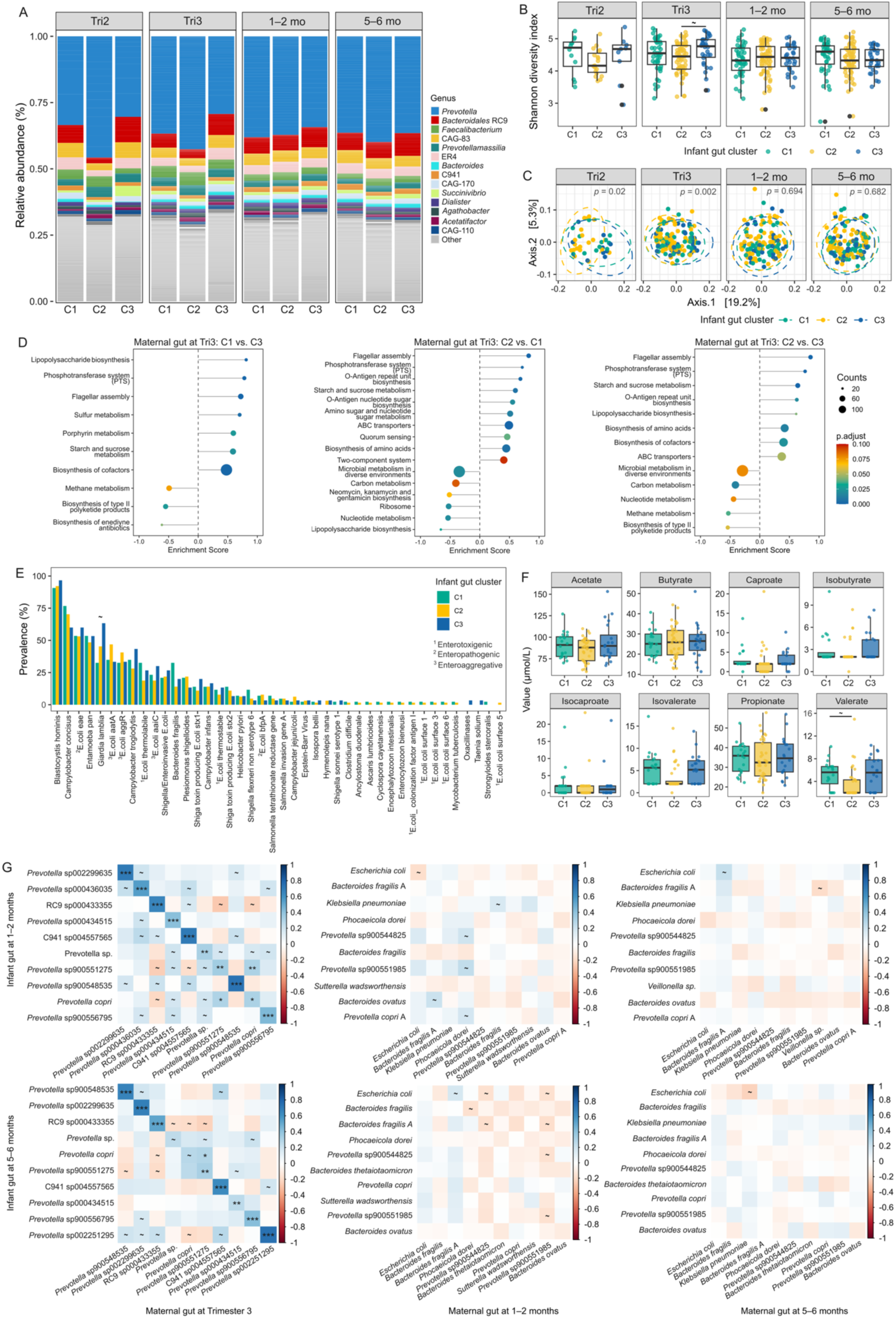
Maternal gut microbiome composition differs across infant gut clusters, particularly during the prenatal period. **A)** Relative abundance of maternal gut genera stratified by infant gut clusters (C1–C3, defined at 1–2 months) across maternal sampling time point: second trimester (Tri2), third trimester (Tri3), 1–2 months, and 5–6 months postpartum. **(B)** Shannon diversity of the maternal gut microbiome by infant cluster and maternal time point. Significance assessed using linear mixed-effects models (LME) with participant as a random effect, followed by pairwise estimated marginal means contrasts. “∼” indicates unadjusted *p* < 0.05 before multiple testing correction (Benjamini–Hochberg). **(C)** Principal Coordinates Analysis (PCoA) of maternal gut microbiome profiles based on Weighted UniFrac distances stratified by infant gut clusters at each time point. Significance was assessed by PERMANOVA with Benjamini–Hochberg correction. **(D)** Gene set enrichment analysis (GSEA) of KEGG pathways in maternal gut metagenomes at the third trimester, comparing mothers of infants in diffrent clusters. Each dot represents a pathway, with size indicating number of contributing genes and color representing FDR-adjusted p-values. **(E)** Prevalence (%) of enteric pathogens detected in the maternal stool at the third trimester, stratified by infant gut cluster. “∼” indicates unadjusted *p* < 0.05 (Ficher’s exact test). **(F)** Short-chain fatty acid (SCFA) concentrations (µmol/L) in maternal stool samples at the third trimester across infant gut clusters. Differences between clusters were assessed using ANOVA. “∼” indicates unadjusted *p* < 0.05 before multiple testing correction. **(G)** Spearman rank correlations between maternal and infant gut taxa shared across ≥10 mother-infant dyads. Correlations matrices are shown for two infant time points and three maternal time points. Color scale represents Spearman’s rank correlation coefficient. \*\*\**p_(FDR)_* < 0.001; \*\**p_(FDR)_* < 0.01; \**p_(FDR)_* < 0.05; ∼ *p*_(unadjusted)_ < 0.05.

Maternal gut microbial diversity, in terms of both alpha and beta diversity, differed across infant clusters during pregnancy. At Tri3, mothers of C3 infants exhibited significantly higher Shannon diversity than C2 (*p_unadjusted_* = 0.04), while no significant differences were observed at other time points (Figure 3B). PCoA further supported these prenatal differences, showing clear compositional separation of maternal gut microbiomes by infant cluster assignment at Tri2 (*p_unadjusted_* = 0.02) and at Tri3 (*p_unadjusted_* = 0.002). This divergence was no longer present postnatally (Figure 3C).

Functional profiling of maternal gut microbiomes revealed distinct metabolic signatures associated with infant gut clusters (Figure 3D). Pathways related to the phosphotransferase system (PTS), starch and sucrose metabolism, and amino acid biosynthesis were enriched in mothers of C2 infants, intermediate in C1, and least represented in C3. Conversely, the lipopolysaccharide (LPS) biosynthesis pathway (often associated with host inflammatory responses) was most enriched in C1 and least in C3. Differences in potential pathogen carriage were also observed across maternal microbiomes. The prevalence of *Giardia lamblia,* a protozoan parasite associated with gastrointestinal disruption, was significantly higher among mothers of C3 infants compared to other clusters (*p_unadjusted_* = 0.03, Figure 3E). Maternal SCFA concentrations were broadly similar across infant clusters, with the exception of valerate levels, which were elevated in mothers of C1 infants compared to C2 at Tri3 (*p_unadjusted_* = 0.04, Figure 3F).

We further evaluated correlations between maternal and infant gut microbiomes within dyads, focusing on shared microbial taxa (Figure 3G). At Tri3, maternal gut taxa correlated positively with the same taxa in the infant gut at both 1–2 months and 5–6 months. Notably, eight of the ten significantly correlated taxa were from the *Prevotella* genus. Postnatally, however, these correlations diminished, and no significant correlations were observed. Interestingly, the composition of shared taxa also shifted, with fewer *Prevotella* species and an increased representation of other taxa after birth.

### Milk microbiome reflects early infant gut clusters and correlates with concurrent infant gut taxa at 1–2 months

Milk microbiome composition was broadly similar across infant gut clusters defined at 1–2 months (Figure 4A). However, milk microbial diversity at later lactation stages appeared to mirror early infant gut clustering. At 3–4 months, mothers of infants in cluster C3 had higher Shannon diversity compared to C1 and C2 (*p_unadjusted_* = 0.02 and 0.03, respectively) (Figure 4B). Beta diversity of the milk microbiome, in contrast, did not differ across infant gut clusters at any time point (Figure 4C).

**Figure 4.**
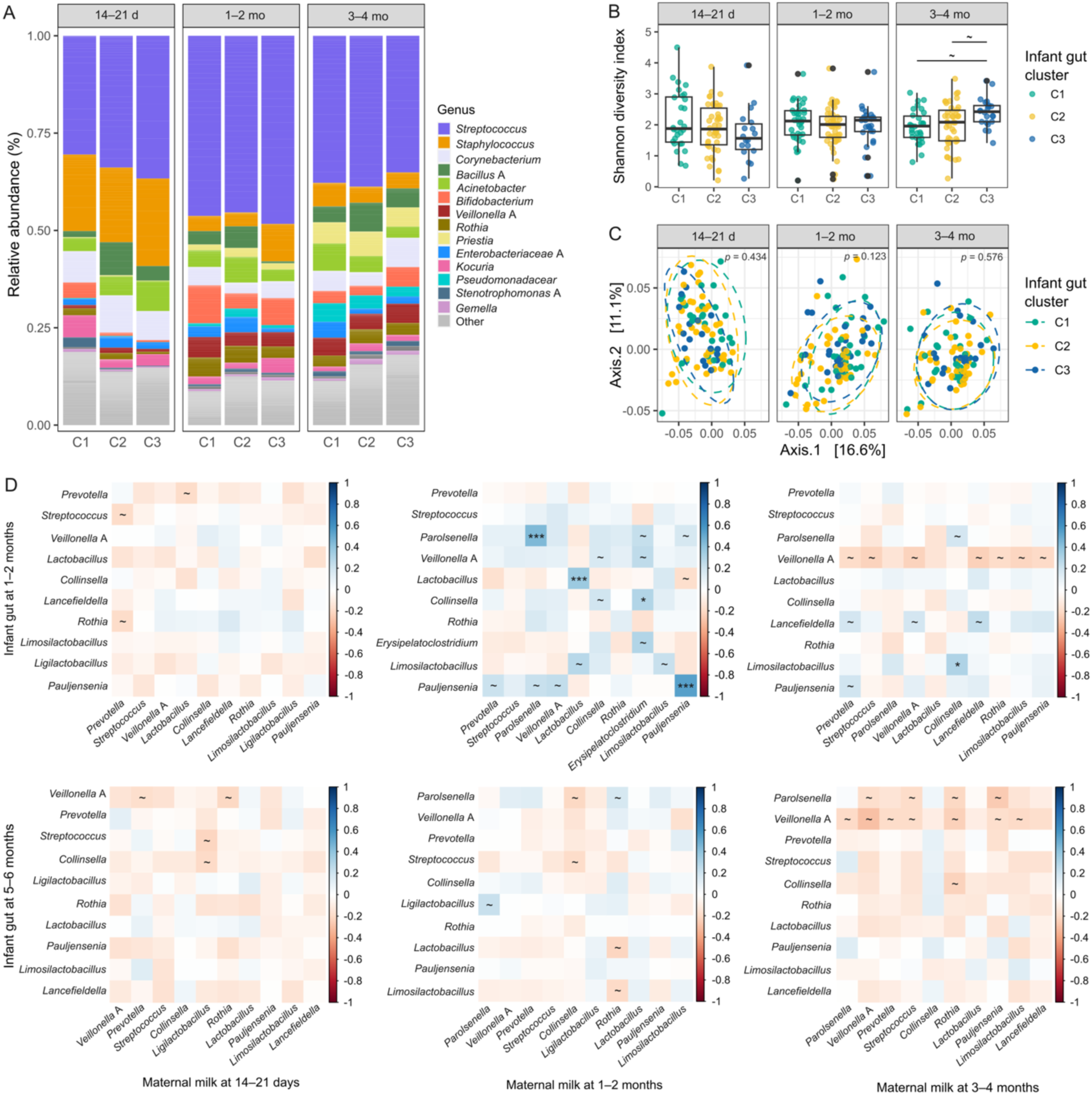
Maternal milk microbiome across infant gut clusters and correlation with infant gut microbiota. **(A)** Relative abundances of the top 15 bacterial genera in maternal milk samples, stratified by infant gut clusters (C1–C3, defined at 1–2 months) and milk sampling time point: 14–21 days, 1–2 months, and 3–4 months postpartum. Genera beyond the top 15 are grouped under “Other.” **(B)** Shannon diversity of the milk microbiome across infant gut clusters and milk sampling time point. Significance was assessed using linear mixed-effects models (LME) with participant as a random effect, followed by pairwise estimated marginal means contrasts. “∼” indicates unadjusted *p* < 0.05 before multiple testing correction (Benjamini–Hochberg). **(C)** Beta diversity of the milk microbiome visualized by Principal Coordinates Analysis (PCoA) based on Weighted UniFrac distances. No significant differences in beta diversity were observed among clusters at any time point. *P*-values were adjusted using the Benjamini–Hochberg method. **(D)** Spearman rank correlations between commonly shared bacterial genera (detected in ≥10 mother–infant dyads) in maternal milk and the infant gut at multiple time points. Top panels show correlations with infant gut microbiota at 1–2 months; bottom panels show correlations with infant gut at 5–6 months. Color scale represents Spearman’s rank correlation coefficient. Significance is indicated after Benjamini–Hochberg correction. \*\*\**p_(FDR)_* < 0.001; \*\**p_(FDR)_* < 0.01; \**p_(FDR)_* < 0.05; ∼ *p*_(unadjusted)_ < 0.05.

We further explored correlations between maternal milk and infant gut microbiomes at postpartum time points. Several milk genera at 1–2 months exhibited significant positive correlations with the same genera in the infant gut at matching time point.

Notably, *Parolsenella, Lactobacillus,* and *Pauljensenia* remained significant after Benjamini-Hochberg correction (Figure 4D). Other genera, such as *Collinsella*, *Erysipelatoclostridium*, and *Limosilactobacillus,* showed initial associations that did not remain significant after correction.

Cross-time correlations revealed additional patterns. Specifically, *Veillonella* abundance in the milk at 3–4 months was negatively correlated with its abundance in the infant gut at 1–2 months, while *Lancefieldella* showed a positive correlation at the same time point. Moreover, *Veillonella* abundance in the infant gut at both 1–2 and 5–6 months was negatively correlated with multiple milk genera measured at 3–4 months. Similarly, the abundance of *Paroisenella* in the infant gut at 5–6 months negatively correlated with several milk genera at 3–4 months.

### HMO composition varies by infant gut cluster and associates with specific gut microbial taxa

HMO secretor status varied across infant gut clusters defined at 1–2 months. The highest proportion was in C2 (88%), followed by C1 (75%) and lowest in C3 (63%), with a statistically significant difference between C2 and C3 (*p* = 0.02, chi-squared test).

Total HMO concentrations were highest in the milk of mothers whose infants were classified into C2, particularly at 14–21 days and 1–2 months (Figure 5A). Several individual HMOs also showed cluster-specific variations. For instance, 2’FL concentrations were significantly higher in the milk of C2 mothers at 14–21 days, whereas lacto-N-tetraose (LNT) were elevated in C3 milk at both 1–2 and 3–4 months. At 3–4 months, 6’-sialyllactose (6’SL), fucosyllacto-N-hexaose (FLNH), and Lacto-N-fucopentaose III (LNFP.III) were also higher in C3, although these differences did not remain significant after Benjamini-Hochberg correction (Figure 5B).

**Figure 5.**
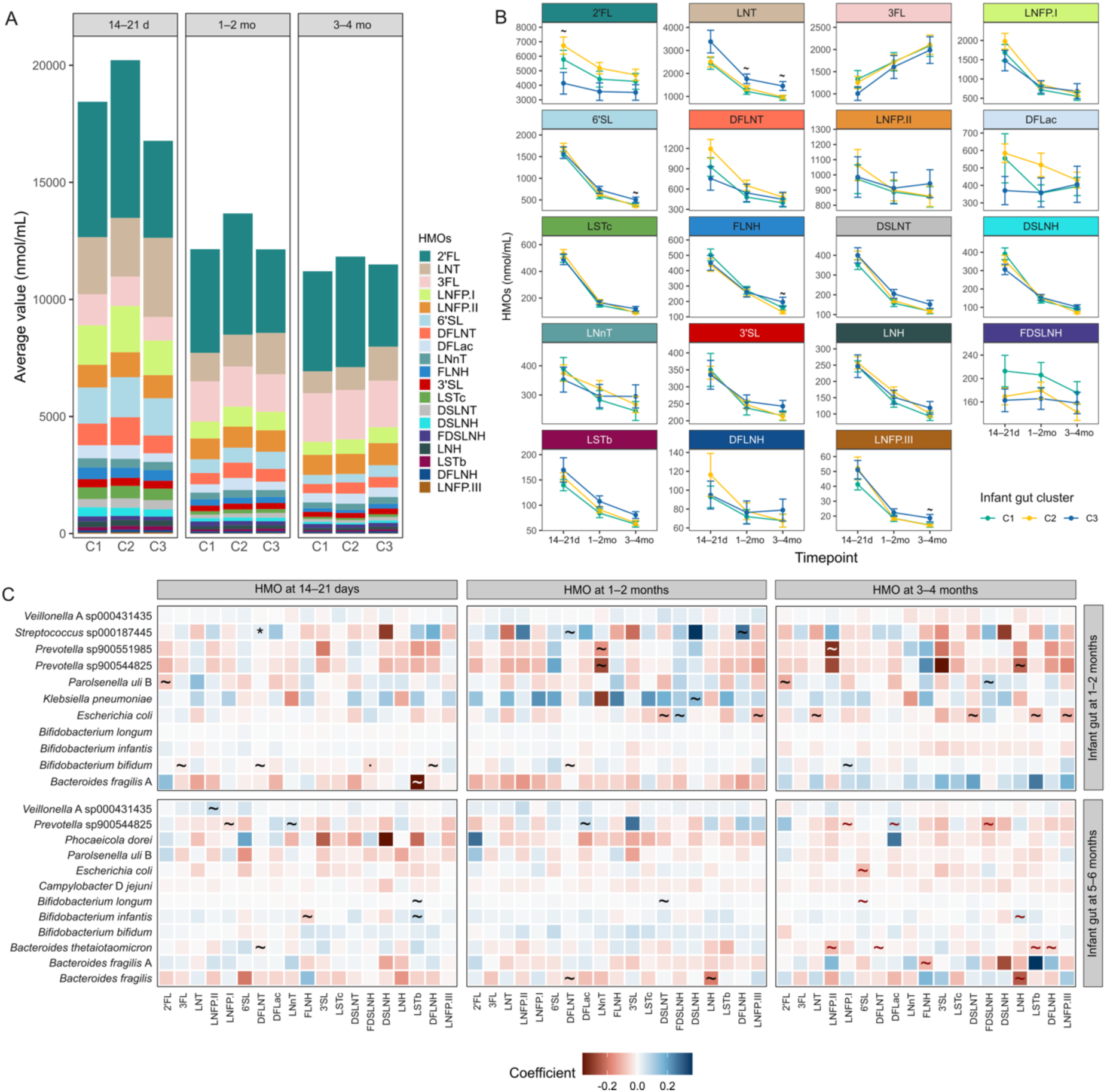
HMO profiles vary by infant gut microbial cluster defined at 1–2 months and associate with specific infant gut taxa. **(A)** HMO concentrations stratified by infant gut cluster (C1–C3, defined at 1–2 months) at three milk sampling time points: 14–21 days, 1–2 months, and 3–4 months postpartum. **(B)** Longitudinal trends of individual HMO concentrations across infant gut clusters. Data shown as means and standard errors. Differences across clusters at each time point were assessed using one-way ANOVA followed by Benjamini–Hochberg correction. “∼” indicates unadjusted *p* < 0.05 before correction. **(C)** Associations between HMO concentrations and the relative abundances of infant gut species (>5% relative abundance) at 1–2 and 5–6 months. Associations were assessed using linear regression models, with HMO levels as independent variables, and secretor status and maternal BEP intervention included as covariates. Tile color reflects the direction and magnitude of the coefficient. \**p_(FDR)_* < 0.05; ∼ *p*_(unadjusted)_ < 0.05.

Associations between milk HMOs and the infant gut species varied over time and were largely taxon-specific (Figure 5C). At 1–2 months, *Streptococcus* sp000187445 showed positive associations with DFLNT across multiple time points, with the association at 14-21 days remaining significant after Benjamini-Hochberg correction (*p*_FDR_ = 0.04). In contrast, *Prevotella* species were negatively associated with several HMOs, such as lacto-N-neotetraose (LNnT). Similarly, *E. coli* was negatively associated with several HMOs, particularly at later time points. *B. bifidum* displayed both positive and negative links across multiple HMOs, with its negative association with fucosyl-disialyllacto-N-hexaose (FDSLNH) remaining a significant trend (*p*_FDR_ = 0.06).

By 5–6 months, fewer positive associations were observed, primarily reflecting early HMO exposures, while negative associations became more common, particularly for *B. fragilis*, and *B. thetaiotaomicron*.

### Milk nutrients and metabolites at 3–4 months reflect 1–2-month infant gut clusters

Distinct nutrient and metabolite profiles in maternal milk at 3–4 months were associated with infant gut clusters defined at 1–2 months. Several milk components, including phosphorus, nicotinamide riboside (NR), and thiamine pyrophosphate (TPP), differed significantly across clusters (*p_FDR_* < 0.05), alongside trends for protein, potassium, arsenic, flavin adenine dinucleotide (FAD), vitamin B2 and NAD (0.05 < *p_FDR_* < 0.1) (Figure 6A). Most of these differences were driven by higher concentrations in cluster C3, which showed elevated levels of fat, protein, energy, phosphorus, potassium, iron, arsenic, and multiple B vitamins at 3–4 months (Figure 6B). In contrast, insulin levels were higher in mothers of C1 infants, and NR levels peaked in mothers of C2 infants.

**Figure 6.**
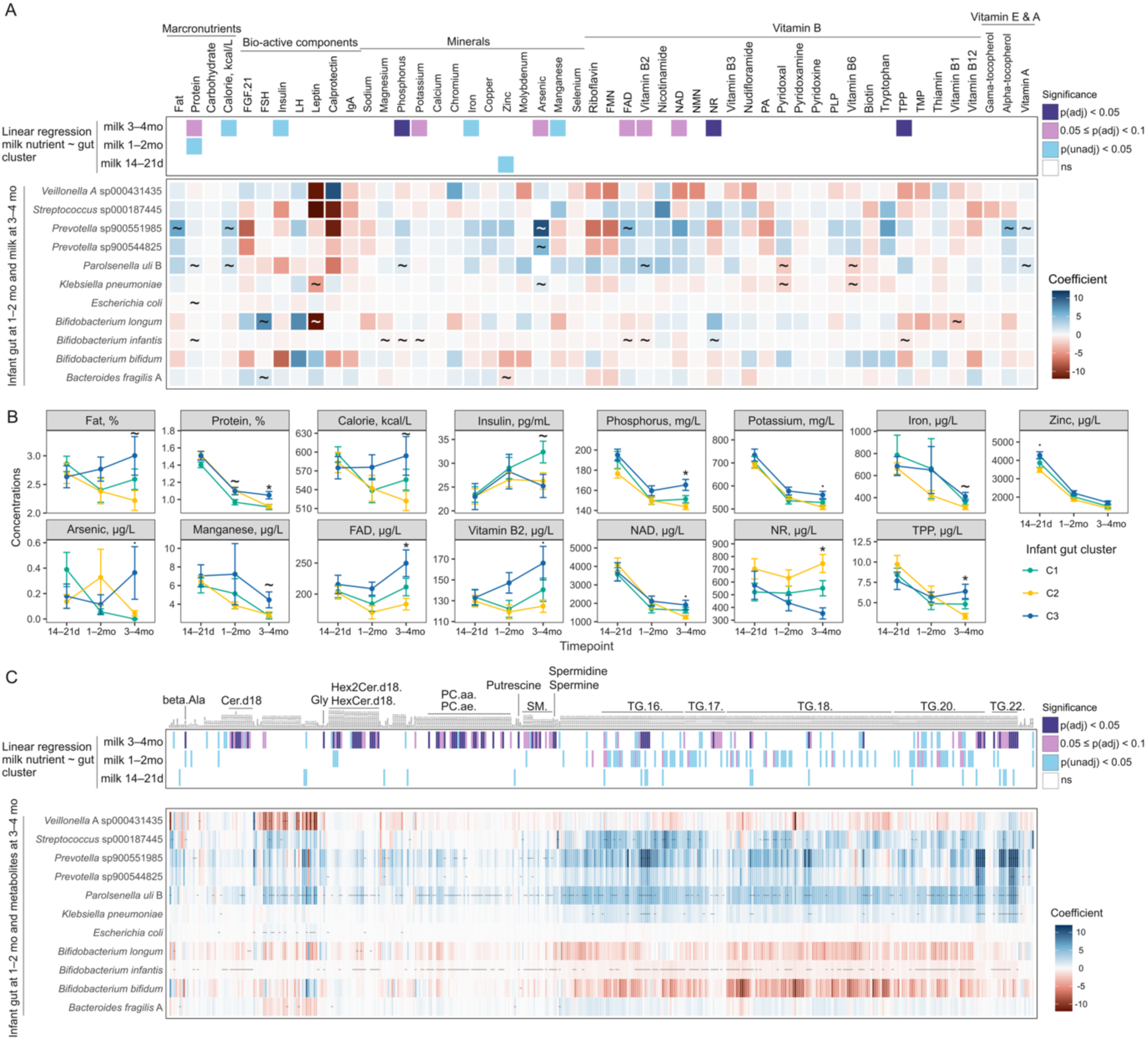
Milk nutrients and metabolites at 3–4 months vary by infant gut clusters defined at 1–2 months and associate with early infant gut microbial composition. **(A)** Top panel: Associations between milk components (macronutrients, bioactive components, minerals, and vitamins) measured at 14–21 days, 1–2 months, 3–4 months, and infant gut clusters (C1–C3 defined at 1–2 months). Associations were assessed using linear regression models with infant cluster as the main predictor and BEP supplementation as a covariate. False discovery rate (FDR) was adjusted by Benjamini–Hochberg correction. Significance is color-coded: dark purple (FDR < 0.05), light purple (0.05 ≤ FDR < 0.1), and blue (unadjusted p < 0.05). Bottom panel: Associations between milk nutrient concentrations at 3–4 months and the relative abundances of dominant infant gut species (>5% abundance) at 1–2 months, based on linear models adjusted for BEP supplementation. Tiles color indicates the regression coefficient. “∼” indicates unadjusted *p* < 0.05 before Benjamini–Hochberg correction. FMN, flavin mononucleotide; FAD, flavin adenine dinucleotide; NAD, nicotinamide adenine dinucleotide; NMN, nicotinamide mononucleotide; NR, nicotinamide riboside; PLP, pyridoxal 5-phosphate; TPP, thiamine pyrophosphate; TMP, thiamine monophosphate. **(B)** Temporal trends and cluster-specific comparisons of milk components (derived from Panel A) across infant gut clusters. Significance assessed using linear regression models with log-transformed nutrient concentrations as dependent variables and cluster as the predictor, and BEP supplementation as a covariate. \**p_(FDR)_* < 0.05; *· 0.05 ≤ p_(FDR)_* < 0.1; ∼ *p*_(unadjusted)_ < 0.05. **C)** Top panel: Associations between milk metabolite concentrations and infant gut clusters, assessed by linear regression models adjusted for BEP supplementation. Each column represents one metabolite, with feature group annotations provided for significant metabolites. Significance color-coded as in Panel A. Bottom panel: Associations between milk metabolites at 3–4 months and infant gut microbial species (>5% abundance) at 1–2 months. Tiles color represents regression coefficient. “∼” indicates unadjusted *p* < 0.05 before multiple testing correction (Benjamini–Hochberg).

We next examined associations between infant gut taxa at 1–2 months and milk components at 3–4 months. *Prevotella* sp900551985 showed positive associations with fat, energy content, arsenic, FAD, alpha-tocopherol and vitamin A. *Parolsenella uli* B was positively associated with protein, energy content, phosphorus, vitamin B2 and vitamin A, while negatively associated with pyridoxal and vitamin B6. *Klebsiella pneumoniae* and *B. longum* were negatively associated with leptin, while *B. longum* and *B. fragilis* A were positively associated with FSH. Notably, *B. infantis* displayed numerous significant associations with milk components, though effect sizes were generally small.

We also compared these findings to associations between infant gut taxa and milk components at the same and later time points, revealing dynamic temporal patterns (Figure S7–S8). For instance, *Prevotella* species at 1–2 months were negatively associated with manganese and pyridoxal 5-phosphate (PLP) in milk collected at the same time, while by 5–6 months, *Prevotella* showed mixed associations with multiple B vitamins measured at 3–4 months. Similarly, *Veillonella* species at 1–2 months were negatively associated with multiple B vitamins (B1, B2, nudifloramine, etc), but by 5–6 months, the strongest associations shifted to zinc and vitamin B3. Together, these results underscore that infant gut–milk relationships are highly time-specific and dynamic.

Finally, metabolomic profiling of milk revealed broad differences in metabolite composition across infant gut clusters, with over half of the 445 analyzed milk metabolites differing significantly by clusters, again most prominently at 3–4 months (Figure 6C). *Streptococcus*, two *Prevotella* species, and *Parolsenella uli* B were consistently associated with higher levels of milk metabolites, particularly triacylglycerols (TGs) containing C16, C17, C18, C20 or C22 fatty acid chains. On the other hand, *Veillonella* and *Bifidobacterium* species, including *B. longum* and *B. bifidum,* were generally negatively associated with these TGs.

### Time-specific and bidirectional associations of maternal milk composition and infant gut microbiome

Redundancy analysis (RDA) revealed that maternal and infant baseline characteristics alone explained a small proportion of the variance in the infant gut microbiome: 3.23% at 1–2 months and 2.50% at 5–6 months. Including maternal gut microbiome, milk microbiome composition, and HMO variables substantially increased the explanatory power of the models. At 1–2 months, the full model accounted for 17.7% of the variance, with 72.5% of this captured by the first two RDA axes (RDA1: 53.7%, RDA2: 18.9%) (Figure 7A). HMOs were the dominant contributors, with concentrations at 14–21 days and 1–2 months explaining 5.4% and 3.8% of the variance, respectively. Shared maternal gut and milk microbiome taxa each contributed approximately 2.4%.

**Figure 7.**
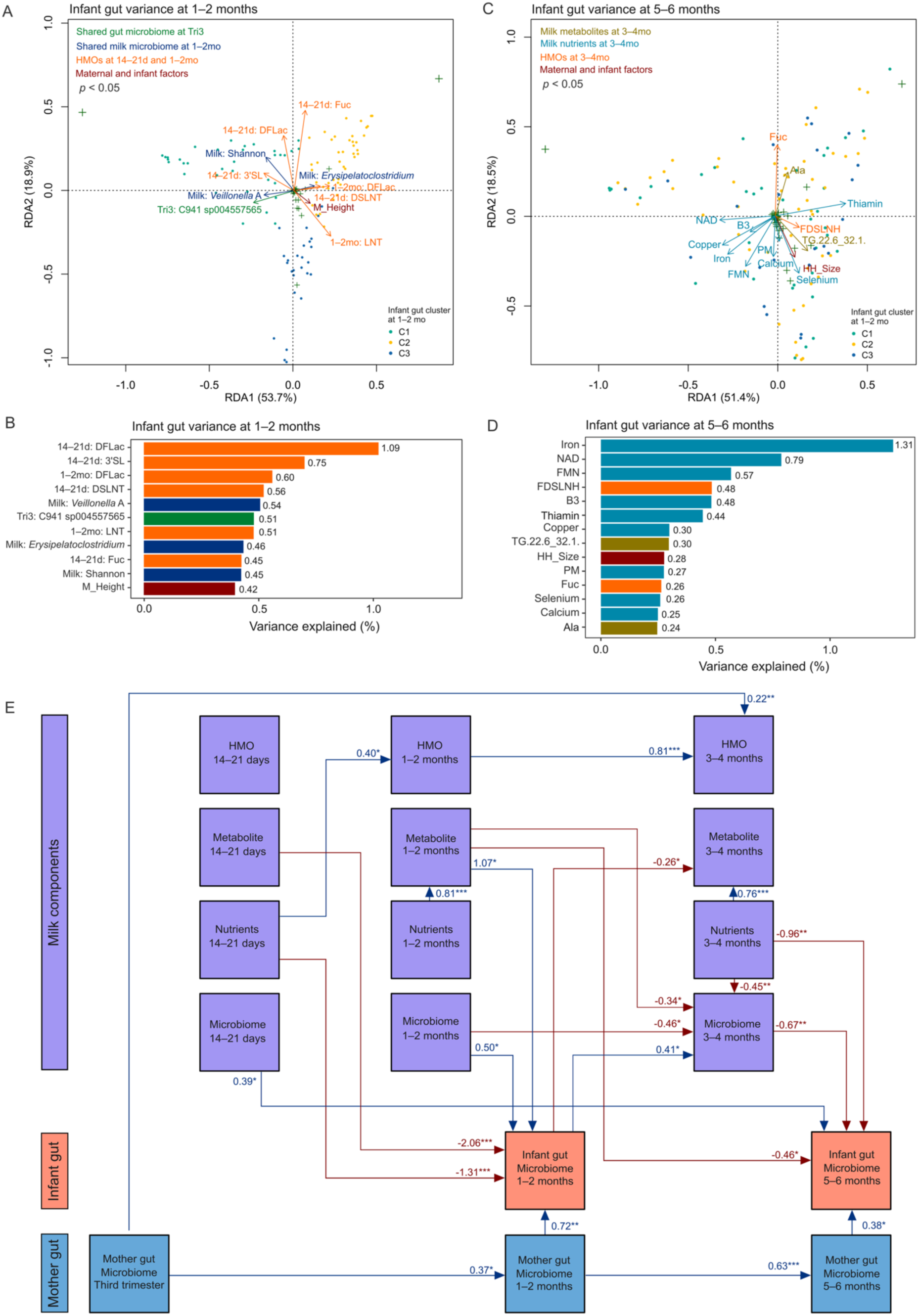
Time-specific and bidirectional associations between milk composition, maternal gut, and infant gut microbiome features. **(A)** Redundancy analysis (RDA) of the infant gut microbiome at 1–2 months including baseline maternal and infant factors (24 features), shared maternal gut taxa and diversity indices from the third trimester (12 features), shared milk taxa and diversity indices at 1–2 months (12 features), and HMO concentrations at14–21 days (21 features), and 1–2 months (21 features). Arrows indicate significant associations (*p* <0.05). Colors represent variable types: green (maternal gut microbiome), blue (milk microbiome), orange (HMOs), and red (baseline characteristics). Full results are provided in Table S4. **(B)** Variance explained by individual significant contributors in the 1–2-month RDA model shown in panel A, grouped and color-coded by feature type. Features with p < 0.05 are shown. **(C)** RDA of the infant gut microbiome at 5–6 months including baseline factors (17 features), milk components (46 features), HMOs (21 features), and the top 15 most important milk metabolites at 3–4 months (selected by XGBoost; see Figure S9). Colors follow Panel A, with teal (nutrients) and gold (metabolites) added. Full results are provided in Table S5. **(D)** Variance explained by individual significant features in the 5–6-month RDA model shown in panel C, grouped and color-coded by feature type. Features with p < 0.05 are shown. **(E)** Structural equation modeling (SEM) based on the first principal components (PC1) scores of each dataset. Arrows indicate directional associations; red indicates negative associations and blue indicates positive associations. Standardized coefficients are shown with significant signs: *p <0.05; **p<0.01; ***p<0.001. Only significant paths (p < 0.05) are displayed for clarity. A complete list of all estimated paths, including non-significant ones, is provided in Table S6.

Significant individual contributors included six HMOs: DFLac (1.09%), 3’SL (0.75%), DSLNT (0.56%), and total fucosylated HMOs (0.45%) at 14–21 days; and DFLac (0.60%) and LNT (0.51%) at 1–2 months. Additional contributors included specific milk and maternal gut taxa, milk Shannon diversity, and maternal height (Figure 7B). While no milk nutrient at 1–2 months significantly explained variance (Table S2), several nutrients measured at 14–21 days, such as protein (0.71%), vitamin B3 (0.70%), and magnesium (0.69), were significant contributors (Table S3).

At 5–6 months, incorporating milk nutrients, HMOs, and the top 15 metabolites identified by XGBoost (based on feature importance; Figure S9) from the 3–4-month time point increased the explained variance to 16.8%, with 69.9% captured by the first two RDA axes (RDA1: 51.4%, RDA2: 18.5%) (Figure 7C). Milk nutrients were the largest contributors (9.42%), surpassing HMOs (2.32%) and milk metabolites (2.09%). Among the 15 significant contributors, nine were milk nutrients, including iron (1.31%), NAD (0.79%), flavin mononucleotide (FMN) (0.57%), vitamin B3 (0.48%), thiamine (0.44%), copper (0.30%), pyridoxamine (0.27%), selenium (0.26%), and calcium (0.25%) (Figure 7F). Other significant contributors included FDSLNH and total fucosylated HMOs, household size and several milk metabolites.

To further explore the directional and temporal structure among maternal gut microbiome, milk composition and infant gut microbiome, we conducted structural equation modeling (SEM) using the first principal component (PC1) scores from each dataset (Figure S10). The final model achieved good fit (CFI = 1.000, RMSEA = 0.000, SRMR = 0.081) and revealed multiple significant associations (Figure 7E). The infant gut microbiome at 1–2 months was strongly influenced by early milk composition, including negative associations with milk metabolites (β = -2.06, *p* < 0.001) and nutrients (β = - 1.31, *p* < 0.001) at 14–21 days, and positive associations with milk microbiome (β = 0.5, *p* < 0.05), metabolites (β = 1.07, *p* < 0.05) and maternal gut microbiome at 1–2 months. In turn, the infant gut at 1–2 months significantly predicted the milk microbiome (β = 0.41, *p* < 0.05) and metabolite (-0.26, *p* < 0.05) at 3–4 months, supporting a bidirectional interaction. At 5–6 months, the infant gut microbiome was negatively associated with milk metabolite (β = -0.46, *p* < 0.05) at 1–2 months, milk nutrients (β = -0.96, *p* < 0.01) and the milk microbiome (β = -0.67, *p* < 0.01) at 3–4 months. Positively associations were observed with the maternal gut at 5–6 months (β = 0.38, *p* < 0.05), and the early milk microbiome at 14–21 days (β = 0.39, *p* < 0.05).

## Discussion

In this comprehensive, longitudinal analysis of milk composition and maternal and infant gut microbiome profiles in a cohort of 152 mother-infant dyads from rural Burkina Faso, we identified three distinct infant gut microbiome clusters at 1–2 months of age, characterized by dominance of *Escherichia*, *Bifidobacterium*, or a more diverse and pathogen-prevalent community. These early-life gut patterns became less distinct by 5–6 months, with increasing overlap in community structure and diversity metrics across infants. Importantly, the clustering patterns not only reflected upstream maternal influences, including the maternal gut microbiota, milk microbiome, and HMO profiles, but were also associated with downstream milk composition, particularly nutrients and metabolites at 3–4 months. In turn, these milk components explained a substantial proportion of the variance in infant gut microbiome composition at 5–6 months.

Together, our findings suggest a time-specific and bidirectional relationship in which early gut colonization is shaped by maternal factors and may in turn influence milk composition, thereby contributing to the trajectory of microbial development in infancy.

In general, the infant gut microbiome undergoes two major transitions: an initial phase after birth during lactation, typically marked by *Bifidobacterium* dominance, followed by a second shift during weaning, when the introduction of solid foods leads to a more diverse, adult-like microbiome dominated by *Bacteroides* and *Firmicutes*^36^.

Bifidobacteria rapidly colonize the infant gut within the first weeks of life, playing a key role in early immune development^37,38^. *E. coli* is another early colonizer, commonly acquired from maternal or environmental sources^39^. While *E. coli* is a typical gut commensal, it also includes multiple pathogenic strains capable of causing both enteric and extra-intestinal diseases^40^. In our study, infants whose gut microbiomes were dominated by *Escherichia* exhibited a higher prevalence of several enteroaggregative *E. coli* strains compared to other clusters, although these differences did not reach statistical significance. Notably, we also observed a subset of infants whose early gut microbiomes were enriched in *Prevotella*, a genus rarely reported in high-income country cohorts^9^, suggesting distinct colonization trajectories in this rural sub-Saharan African setting.

In our study, infants with a more diverse gut microbiome early in life (Cluster C3) exhibited higher levels of certain SCFAs at 1–2 months, alongside a greater prevalence of enteric pathogens such as *Salmonella* and *Adenovirus*, suggesting broader microbial exposures and potentially reflecting an environmentally influenced or adult-like community structure^41,42^. An epidemiological study found that *Salmonella* infection in infants remains a significant cause of morbidity and mortality in the U.S., particularly among younger infants and those identified as Black or Asian (based on parent-reported race/ethnicity), with routes of exposure potentially varying by environmental and behavioral factors^43^. *Adenoviruses*, particularly serotypes 40 and 41, are common causes of gastrointestinal infections, diarrhea, and gastroenteritis in infants and young children^44^. Unlike infants in clusters C1 and C2, who showed rising SCFA concentrations over time, C3 infants exhibited a decline in total SCFA levels from 1–2 months to 5–6 months, suggesting that peak metabolic activity may have occurred earlier. This pattern may indicate accelerated microbial maturation in C3 infants.

SCFAs, primarily produced through the fermentation of non-digestible carbohydrates, are generally beneficial to host metabolic and immune health^45–49^. Protein fermentation may also contribute, with amino acid metabolism yielding SCFAs such as acetate and propionate^47^, while isovalerate is typically associated with proteolysis and gut protein breakdown^45,50^. Interestingly, in our study, mothers of infants in the more diverse gut cluster (C3) exhibited higher protein concentrations in their milk, suggesting a potential link between milk nutrient provision and infant gut microbial metabolic activity.

The origins of early microbial composition remain multifactorial. While maternal and infant baseline characteristics were broadly comparable across infant gut clusters in our study, we observed trends toward higher gestational weight gain and a greater number of children under five in the household among infants in the *Bifidobacterium*-dominated group. These differences raise the hypothesis that subtle variations in maternal metabolic status and family structure may influence early microbial development. In particular, a higher number of young children in the household may reflect ongoing breastfeeding of multiple children, potentially increasing opportunities for *Bifidobacterium* transmission from both the mother and milk-fed siblings to the focal infant. Previous studies have reported that increased parity and household crowding are associated with altered microbial transmission dynamics and early-life exposures to a broader microbial pool^13,51^. Similarly, gestational weight gain may reflect maternal nutritional status and metabolic state, which can influence fetal development and potentially shape microbial environments^52–54^.

By 5–6 months, the infant gut microbiome in this study was largely characterized by the dominance of *Bifidobacterium* and *Escherichia*. This transition likely reflects shared environmental exposures and feeding practices. In our cohort, nearly all infants were exclusively breastfed, with average durations close to six months—a factor known to promote a *Bifidobacterium*-rich gut environment through the provision of HMOs, antimicrobial peptides, and other bioactive components^55,56^. This strong breastfeeding pattern may have acted as a unifying selective pressure, shaping the infant gut microbiome toward a common microbial profile across infants.

A key finding from the present analysis is the temporally distinct and stage-specific roles of maternal and milk-derived factors on infant gut microbiome development. During early infancy (1–2 months), the maternal gut microbiome, particularly higher prenatal *Prevotella* abundance, along with early HMO profiles, emerged as the dominant predictors of infant gut microbial variation. Specific HMOs, including DFLac, 3’SL and LNT are well-known to function as selective substrates for beneficial *Bifidobacterium* and *Bacteroides* species^18^, providing a substrate-driven mechanism for vertical microbial transmission^57,58^. In contrast, at 5–6 months, HMOs explained a smaller proportion of the variation, while milk nutrients and metabolites, especially iron and several B vitamins measured at 3–4 months, became the primary explanatory factors. This temporal shift from early microbiome structuring via maternal-derived selective substrates to later modulation by the broader nutritional landscape of milk, offers new insight into how different components of milk contribute to microbial succession. Many of these nutrients have known roles in microbial ecology: for example, iron availability is tightly regulated in the gut, with lactoferrin in human milk sequestering iron to limit its accessibility to pathogenic bacteria^59,60^. Similarly, vitamin B12 is not only essential for the host but also for many microbial metabolic pathways, influencing species composition and competition within the gut^61^. Together, these findings underscore the time-resolved influence of milk composition on the establishment and maturation of the infant gut microbiome.

The present findings suggest potential feedback loops whereby the infant gut microbiome may influence maternal milk composition. Infants with more diverse early microbiomes were associated with higher levels of macronutrients, essential minerals, B vitamins, and a wide range of milk metabolites at 3–4 months postpartum. While causality cannot be established in this observational setting, our SEM analysis supports the directionality of this association, specifically that variation in the infant gut microbiome at 1–2 months significantly predicts milk microbiome composition and metabolites at 3–4 months. These patterns align with emerging evidence that maternal milk composition is dynamic and may respond to infant-derived cues through several plausible biological pathways^24,62^. One possibility is that microbial metabolites produced in the infant gut (such as SCFAs or other signaling molecules) enter systemic circulation and influence salivary composition^63,64^, thereby delivering signals to the maternal breast during suckling^24^. Alternatively, the infant oral microbiome, which share taxa and have bidirectional associations with the gut microbiome^65^, may be transferred to the mammary gland through retrograde flow during nursing, directly impacting the milk microbiome^66,67^ and potentially influencing milk biosynthesis. In addition, variation in infant gut microbial communities could impact immune development and cytokine expression, which may trigger maternal immune response and alter the composition of immunomodulatory or metabolic components in milk.

Supporting this, recent studies have shown that infant infections can lead to rapid increases in immunomodulatory constituents in breast milk, which subside upon infant recovery^29,68^. Together, these observations support the intriguing possibility that maternal-infant dyads engage in adaptive, bidirectional communication, enabling mothers to fine-tune milk composition in response to the infant’s developmental needs and health status.

Key strengths of this study include its time-resolved multi-omic design, which allowed for integrated analyses of maternal gut, milk composition, and infant gut microbiomes across multiple time points in a low-resource setting. The inclusion of rare, high-resolution multi-omic data from 152 breastfeeding mother-infant dyads in rural Burkina Faso represents a significant contribution, given the persistent underrepresentation of LMIC populations in microbiome research. By combining shotgun metagenomics, 16S sequencing, HMO quantification, pathogen detection, and nutrient/metabolite assays, this study offers a comprehensive view of microbial and nutritional dynamics during early life. Although the analysis presented here is observational and exploratory, it benefits from the underlying cohort being drawn from a randomized controlled trial context, which minimized selection bias and exposure heterogeneity. Finally, while the generalizability of our findings may be bounded by this specific low-resource, breastfeeding-predominant context, this setting offers unique scientific advantages. In particular, the extended duration of exclusive breastfeeding provided a valuable opportunity to explore the temporally dynamic associations between milk composition and infant gut microbiome development, an aspect often difficult to assess in cohorts from high-income settings where early weaning and mixed feeding are more common.

Nonetheless, several limitations should be considered. While we measured hygiene access and infant feeding practices, we lacked direct biomarkers of systemic inflammation, which could offer additional context for interpreting pathogen and SCFA profiles. Our observational framework also limits causal inference, and while some maternal and infant characteristics differed by cluster, these associations were modest and should be interpreted cautiously. Additionally, the use of 16S rRNA gene sequencing for milk samples, due to the low microbial biomass and high host DNA content in human milk, limited taxonomic resolution compared to metagenomic sequencing and hindered strain-level tracking of maternal-infant microbial transmission through milk.

Taken together, our findings highlight a complex, temporally coordinated landscape of maternal–infant interactions that shape early-life gut microbiome trajectories. Maternal gut and milk microbiomes, alongside dynamic shifts in milk oligosaccharides, nutrients and metabolites, were linked to infant gut microbial development. Importantly, early gut profiles were also associated with later milk composition, supporting the possibility of adaptive, bidirectional communication between maternal physiology and infant needs. These insights emphasize the importance of maternal prenatal health and lactational biology in early-life microbial development and provide a foundation for future work to unravel mechanistic pathways and inform microbiome-targeted interventions.

## STAR Methods

### KEY RESOURCE TABLE

| RESOURCE | SOURCE | IDENTIFIER |
| --- | --- | --- |
| Biological samples |  |  |
| Breastmilk | Bastos-Moreira et al., 2023<sup>33</sup> | NA |
| Stool | Bastos-Moreira et al., 2023<sup>33</sup> | NA |
| Deposited Data |  |  |
| Raw metagenomic sequenced data | Deng et al., 2025<sup>34</sup> | NCBI database under BioProject accession code PRJNA1113165 |
| Raw 16S rRNA sequenced data |  | NCBI database under BioProject accession code PRJNA1292286 |
| Software and packages |  |  |
| R (v4.3.1) | R Core Team | https://www.r-project.org |
| phyloseq (v1.44.0) | McMurdie and Holmes, 2013<sup>69</sup> | https://joey711.github.io/phyloseq |
| vegan (v2.6-4) | Oksanen et al., 2001<sup>70</sup> | https://cran.r-project.org/package=vegan |
| ANCOM-BC2 (v2.2.1) | Lin and Peddada, 2020<sup>71</sup> | https://github.com/frederickhuanglin/ANCOMBC |
| treeclimbR (v0.1.5) | Huang et al., 2021<sup>72</sup> | https://bioconductor.org/packages/treeclimbR |
| edgeR (v4) | Chen et al., 2025<sup>73</sup> | https://bioconductor.org/packages/edgeR |
| xgboost (v1.7.4) | Chen and Cuestrin, 2016<sup>74</sup> | https://xgboost.readthedocs.io/en/release_3.0.0/ |
| Lavaan (v0.6-19) | Rosseel et al, 2012<sup>75</sup> | https://github.com/yrosseel/lavaan |

## RESOURCE AVAILABILITY

### Lead contact

Further requests for information should be directed to and will be fulfilled by the Lead Contacts, Trenton Dailey-Chwalibóg and Carl Lachat

### Material availability

This study did not generate new unique reagents.

### Data and code availability

The metagenomic sequencing data generated in this study have been deposited in the NCBI database under BioProject accession code PRJNA1113165 [https://www.ncbi.nlm.nih.gov/bioproject/PRJNA1113165]. The processed taxonomy and gene data are available at https://github.com/stefftaelman/misame-metagenomics-BEP.

The accession numbers for the 16S rRNA gene sequence data reported in this paper are BioProject accession (NCBI): PRJNA1292286. The other milk data can be accessed upon request to Meghan Azad (meghan.azad@ umanitoba.ca)

The de-identified personal data are available under restricted access for the purpose of verifying, interpreting, or extending the research published in this article. Access can be obtained by contacting and signing a data-sharing agreement. A response to requests for data access can be expected within 2 months, and the data will be available for one year after signing the data-sharing agreement.

## EXPERIMENTAL MODE AND SUBJECT DETAILS

### Study design and cohort description

This study was nested within the MISAME-III randomized controlled trial, conducted in the Houndé district of the Hauts-Bassins region in rural Burkina Faso. The full study protocol has been published previously^30^. MISAME-III was an individually randomized 2×2 factorial trial designed to evaluate the efficacy of BEP supplementation, provided to women during pregnancy and the first six months of lactation, on small-for-gestational age prevalence and length-for-age z-scores at six months^31,32^. Women were eligible if they had a confirmed pregnancy by urinary and ultrasound tests and provided written informed consent. Excluded criteria included gestational age ≥21 weeks, peanut allergy, or plans to relocate. The study enrolled 1,879 pregnant women aged 15-40 years from October 30, 2019, to December 12, 2020. Women were randomized to receive either BEP supplementation with iron and folic acid tablets (i.e., prenatal intervention group) or iron and folic acid tablets alone (i.e., prenatal control group) during pregnancy and again randomized at delivery for postnatal intervention and control groups. The BEP supplement was a lipid-based, ready-to-use peanut paste enriched with multiple micronutrients and manufactured by Nutriset. A daily 72 g sachet delivered 393 kcal (36% from fat, 20% from protein, 32% from carbohydrates), with protein sourced from soy, milk, and peanuts. The formulation covered estimated average requirements for 11 essential micronutrients, though not for calcium, magnesium, or phosphorus. All women received IFA tablets (65 mg iron, 400 µg folic acid) as part of standard prenatal care, along with malaria prophylaxis per national guidelines (sulfadoxine–pyrimethamine).

The biospecimen sub-study (Biospé) was initiated after the main MISAME-III trial recruitment was completed and aimed to collect biospecimens from a subset of mother-infant dyads to evaluate the physiological effects of the supplementation on women and their infants^33^. At the time of its launch, most participants were already in their third trimester; therefore, to enrich the cohort with women in their second trimester, recruitment prioritized individuals with earlier gestational age, ensuring even distribution across all four study groups. A total of 309 mother-infant dyads were enrolled in Biospé, of whom 152 dyads were selected for metagenomic sequencing due to budget constraints: 71 in the pre-and postnatal intervention group, and 81 in the pre-and postnatal control group. Milk microbiome and profiling for other components were done on the full subset, but in this analysis, we only focused on the 152 dyads which allow us to integrate with the gut metagenomic data.

Maternal stool samples were collected at four time points: second trimester (19–24 weeks), third trimester (30–34 weeks), 1-2 months postpartum (25–56 days), and 5-6 months postpartum (147–175 days). Infant fecal samples were collected at 1–2 months and 5–6 months after birth, aligned with maternal sampling. Human milk samples were collected at 14–21 days, 1–2 months (28–56 days) and 3–4 months (84–112 days) postpartum. A total of 472 maternal stool, 423 milk, and 256 infant stool samples were included in the analysis.

Maternal and infant anthropometric measurements, household and maternal baseline characteristics, feeding practices, and clinical assessments were collected by study physicians and midwives using SurveySolution (version 21.5) on tablets.

### Ethical approvals

The study was approved by the Commissie voor Medische Ethiek (CME) of Ghent University Hospital (protocol code: B670201734334 and date of 10/08/2020) and the Comité d’Éthique Institutionnel de la Recherche En Sciences de la Santé (CEIRES) of the Institut de Recherche en Sciences de la Santé (IRSS) (protocol code: 50-2020/ CEIRES and date of 22/10/2020). An independent Data and Safety Monitoring Board (DSMB), which included an endocrinologist, two pediatricians, a gynecologist, and an ethicist from Belgium and Burkina Faso, was established prior to the trial. The DSMB managed remote safety reviews for adverse and serious events at nine and 20 months after the start of enrollment. The MISAME-III trial was registered on ClinicalTrials.gov (identifier: NCT03533712).

## METHOD DETAILS

### Sample collection

Maternal fecal samples (∼8 g) were collected using sterile stool collection containers, aliquoted into sterile cryotubes (Biosigma, Cona, VE, Italy), and immediately flash-frozen before long-term storage at −80 °C. Infant feces (∼8 g) were collected using a 38 × 50 cm sterile protection sheet (Kimberley-Clark, Irving, TX, USA), wrapped around the infant like a diaper, then transferred to OMNIgene•GUT OM-200 collection kits and stored at−80 °C. Fecal consistency was assessed using the visual Bristol scale. Liquid samples were homogenized to ensure uniformity before aliquoting. During transport, samples were kept on dry ice with continuous temperature logging and subsequently stored at −80 °C until analysis.

Breastmilk was collected using an electric breast pump (Medela, Baar, Switzerland) from the breast opposite the one most recently used for feeding. A full expression was performed, and 7.2 mL of milk was collected and aliquoted into four 2 mL sterile cryotubes. Samples were gently inverted to ensure mixing of fore- and hindmilk.

### Metagenomic sequencing for stool samples

DNA extraction, library preparation, and metagenomic sequencing were performed at the University of California San Diego IGM Genomics Center. DNA was extracted using the Thermo MagMAX Microbiome Ultra kit and quantified using the PicoGreen assay. Library preparation was conducted with Roche KAPA HyperPlus kits using 25 ng of DNA input and 9 PCR cycles. Shallow MiSeq Nano sequencing was used to normalize read sequencing depth before high-throughput paired-end 2 × 150 bp sequencing on an Illumina NovaSeq 6000 at a target depth of 25 Gbp/sample for maternal samples and 10 Gbp/sample for infant samples.

Reads were trimmed and deduplicated using FastP, and human reads were removed via alignment to the hg38 reference genome using Bowtie2. Microbial reads were mapped to the DeltaI reference genome database, optimized for profiling microbiomes from non-industrialized infants^9^. Relative abundances of genomes were calculated as the proportion of mapped reads per genome to total non-human reads. Only genomes with >50% breath coverage were retained. Functional gene annotation was performed using the inStrain pipeline under default parameters^76^.

### Pathogen detection in stool

Enteric pathogens were screened using a customized read-time PCR-based TaqMan Array Card (TAC) with real-time qPCR assays targeting >80 targets, including genomic targets from bacteria, viruses, *Shigella* speciation and serotyping targets, colonization factors of enterotoxigenic *Escherichia coli*, and gene targets associated with antimicrobial resistance^77^. Total nucleic acids were extracted from stool samples using a modified QIAamp Fast DNA Stool Mini Kit with bead beating and heat incubation to enhance yield. Pathogen burden was determined by quantification cycle (Cq), and a Cq value <35 was considered positive.

### Short-chain fatty acid analysis in stool

Short-chain fatty acid (SCFAs) and branched-SCFAs were quantified from stool samples at ProDigest (Belgium). Samples were partially freeze-dried to determine water content, and extraction was performed followed by gas chromatography with flame ionization detector. The following compounds were analyzed: acetate, propionate, butyrate, isobutyrate, isovalerate, valerate, isocaproate, and caproate. Concentrations were reported in µmol/g of wet feces. Values reported as “ND” (not detected) were treated as zero, while values below the limit of quantification (“<LOQ”) were imputed as half of the corresponding LOQ value.

### Milk microbiota profiling

Milk microbiota was analyzed at Baylor College of Medicine by 16S rRNA gene sequencing of the V4 hypervariable region as previously described^78,79^. Milk samples were preprocessed for this analysis with consideration for its low biomass, including negative controls to assess contamination in all processes, including initial aliquoting. DNA was extracted from 1 mL of milk after centrifugation (10,000 ×g) for 5 minutes, using the DNeasy 96 PowerSoil Pro HT Kit and Qiagen QIACube HT automated extraction platform. Contaminants were identified and removed using Decontam^80^ and SCruB algorithms^81^ with negative controls included at each step.

### Other milk components

Human milk samples were analyzed for HMOs, macronutrients, micronutrients, bioactive proteins and metabolites using established protocols as previously described^82^.

HMOs were quantified at the Mother-Milk-Infant Center of Research Excellence (MOMI CORE) at the University of California, San Diego. HMOs were isolated from milk using high-throughput solid-phase extraction, fluorescently labeled, and analyzed by high-performance liquid chromatography with fluorescence detection (HPLC-FLD) as described previously^83,84^. Identification and quantification of HMOs were based on retention times and mass spectrometry, using raffinose as an internal standard for absolute quantification.

Macronutrients, including fat, protein, carbohydrate (mainly lactose) and energy content, were measured by near infrared spectroscopy (NIR) using a SpectraStar XT (KPM analytics, Westborough, MA, USA) calibrated specifically for human milk, as previously described^85,86^. These analyses were performed at the Manitoba Interdisciplinary Lactation Center (MILC) biorepository in Canada.

Micronutrients were analyzed at the United States Department of Agriculture (USDA) Agricultural Research Service (ARS)-Western Human Nutrition Research Center (WHNRC) in Davis, CA, USA using the same methods described for the Mothers, Infants and Lactation Quality study^87^. This included multiple B-vitamins^88–90^, fat-soluble vitamins (vitamins A and E)^91^, and minerals and trace-elements (calcium, copper, iron, magnesium, potassium, selenium, sodium, and zinc)^92^. Choline, a water-soluble compound that plays roles in common metabolic pathways with B-vitamins, was measured by liquid chromatography-tandem mass spectrometry (LC-MS/MS, Biocrates Life Science, Austria) as part of the metabolomics analysis described below^93^.

Bioactive proteins in milk were quantified using high-performance electrochemiluminescence assay (Meso Scale Discovery, USA at MOMI CORE)^94^. These proteins were selected for their immunological and metabolic relevance in infant health and included secretory Immunoglobulin A (sIgA), calprotectin, and metabolic hormones such as FSH, luteinizing hormone, insulin, leptin, and fibroblast growth factor-21 (FGF-21). Assays were multiplexed where concentration ranges allowed, and assay conditions were optimized for human milk matrices.

Targeted milk metabolomics profiling was performed at Biocrates Life Science (Innsbruck, Austria) using LC-MS/MS and flow injection analysis-tandem mass spectrometry (FIA-MS/MS) with the MxP Quant 500 kit^87,93,95^. This validated platform allowed quantification (µmol/L) of over 600 known metabolites, including 242 triglycerides, 12 free fatty acids, 20 free amino acids, and 40 acylcarnitines, providing broad coverage of lipid, amino acid, and energy metabolism.

## QUANTIFICATION AND STATISTICAL ANALYSIS

To account for its potential confounding effects, all association analyses in the present study were adjusted for intervention status.

### Clustering analysis

Clustering analysis was performed using genus-level relative abundances of the infant gut microbiome at 1–2 months of age, and processed using the phyloseq (v1.44.0) R package. Taxonomic profiles were aggregated at the genus level, and the top 15 most abundant genera across all samples were retained for clustering, while less abundant genera were grouped as “Other” for interpretability. Hierarchical clustering was performed using Bray-Curtis dissimilarity and Ward’s minimum variance method (vegan v2.6-4). The optimal number of clusters was determined by evaluating clustering metrics, including silhouette width, Calinski-Harabasz index, and within-group dispersion. Three clusters were selected based on these metrics and biological interpretability, corresponding to dominant genera including *Escherichia*, *Bifidobacterium*, and a diverse profile enriched in *Bacteroides* and *Prevotella* species. Cluster membership was visualized using heatmaps, and subsequent longitudinal analyses were performed to assess cluster convergence across later time points.

### Diversity analysis

Alpha and beta diversity analyses were performed for the infant gut microbiome, maternal gut microbiome, and milk microbiome using the phyloseq (v1.44.0) and vegan (v2.6-4) R packages.

Alpha diversity metrics, including Shannon diversity index and observed richness, were calculated based on rarefied count data. For pairwise comparisons of alpha diversity across infant gut microbiome clusters at each time point, linear mixed-effects models were fitted using the nlme package^96^, with cluster as the fixed effect and participant ID as a random intercept. Pairwise contrasts were conducted using emmeans (estimated marginal means), and p-values were adjusted for multiple testing using the Benjamini-Hochberg false discovery rate (FDR) correction.

Beta diversity was assessed using weighted UniFrac distance metrices, calculated from phylogenetic trees within phyloseq (v1.44.0). Principal Coordinate Analysis (PCoA) was used for ordination visualization, and 95% confidence ellipses were drawn for each cluster. Permutational multivariate analysis of variance (PERMANOVA) was conducted using adonis2 from the vegan package to test for significant differences in community composition between clusters and over time. P-values from PERMANOVA tests were adjusted for FDR.

### Differential abundance analysis

Differential abundance analysis of microbial taxa was performed at the species level using the ANCOM-BC2 (v2.2.1) framework implemented in R. Analyses were conducted separately for the infant gut microbiome at 1–2 months to identify taxa differentially abundant among the three clusters identified from the clustering analysis.

Raw count data from phyloseq objects were used as input, and low-prevalence taxa (present in <10% of samples) and samples with sequencing depth <1000 reads were excluded. ANCOM-BC2 models were fitted with cluster as the fixed effect, without random effects, using the default settings for iterative convergence, empirical Bayes shrinkage, and bias correction. Structural zeros were identified and accounted for in the model. The Benjamini-Hochberg method was applied to control the FDR. Pairwise differential abundance comparisons were performed between clusters (C2 vs. C1, C2 vs. C3, and C1 vs. C3) using the ANCOM-BC2 pairwise contrast framework. Log2 fold-changes and FDR-adjusted p-values were calculated for each comparison.

### Multivariate models

To evaluate the combined effects of maternal, milk, and infant factors on the infant gut microbiome composition, redundancy analysis (RDA) was performed using the vegan package (v2.6-4) in R.

Infant gut microbiome composition, based on species-level relative abundance profiles, at 1–2 months and 5–6 months was used as the response matrix. Taxa detected in fewer than 10% of samples were excluded to reduce sparsity and limit the influence of rare species.

For the RDA model of the infant gut microbiome at 1–2 months, we constructed a comprehensive covariate matrix incorporating infant characteristics (birth weight, length, head circumference, mid-upper arm circumference, gestational age at birth, and duration of exclusive breastfeeding), maternal characteristics (maternal age, body mass index, weight, height, mid-upper arm circumference, hemoglobin concentration, dietary diversity score, gestational weight gain, parity, and BEP intervention status), and household factors (household size, number of children under five years, household food insecurity, lean season during sampling, and reported infant “purge” practice). Additionally, we included the shared maternal gut microbiome during the third trimester, the shared milk microbiome at 1–2 months, and HMO profiles at 14–21 days and 1–2 months.

For the RDA model of the infant gut microbiome at 5–6 months, the covariate matrix similarly included maternal, infant, and household characteristics, as well as milk composition profiles at 3–4 months. Milk composition data comprised macronutrients, micronutrients, bioactive proteins, HMOs, and key milk metabolites identified through machine learning models (described below).

The significance of the global RDA model and individual covariates was assessed using permutation tests with 999 permutations. The proportion of variance explained by the model and by each covariate was calculated based on adjusted R² values.

### Machine learning models

To identify important milk metabolites associated with infant gut microbiome clustering at 1–2 months, we applied a supervised machine learning approach using extreme gradient boosting (XGBoost)^74^ classification models implemented in the xgboost R package (v1.7.4).

Infant gut microbiome clusters (derived from hierarchical clustering, described above) were used as the classification outcome. Milk metabolite profiles at 3–4 months were used as the feature matrix, after excluding variables with low variance or missingness. The dataset was randomly split into training (75%) and testing (25%) subsets.

Model training was performed on the training set using five-fold cross-validation to optimize the number of boosting rounds based on the minimum multi-class log-loss error. The final model was trained using the optimal number of rounds and evaluated on the independent testing set.

Classification performance was assessed using accuracy, confusion matrices, and area under the receiver operating characteristic curve (AUC) calculated for each cluster. Variable importance was ranked based on model-derived gain scores. Given that the final model achieved an AUC of 0.687, top-ranked metabolites were further explored in association analyses with infant gut microbial species abundances.

### Structural equation modeling

Structural equation modeling (SEM)^97^ was used to investigate longitudinal and bidirectional relationships among maternal gut microbiome, milk composition (microbiome, metabolite, nutrients, and HMOs), and infant gut microbiome across sampling time points. To capture the variation of each dataset, the first principal component (PC1) scores from each dataset were used in the SEM. Variables were scaled prior to modeling. For microbial datasets, we first computed weighted UniFrac distances and then performed Principal Coordinates Analysis (PCoA). The first PCoA axis representing the greatest variance in community composition, was extracted as the PC1 score. For other milk components, PC1 scores were derived using Principal Component Analysis (PCA) and the first PC was extracted. SEM models were fitted using the lavaan package (v 0.6-19)^75^. An initial path model was iteratively refined using modification indices, retaining only significant relationships while monitoring model fit criteria: comparative fit index (CFI) >0.95, root-mean-square error of approximation (RMSEA) <0.05 and standardized root-mean residuals (SRMR) <0.08^98^. This data-driven approach led to a final SEM with good model fit (CFI = 1.000, RMSEA = 0.000, SRMR = 0.078), capturing time-resolved direct and indirect influences from maternal and milk factors to the infant gut microbiome, as well as feedback pathways from infant gut status to subsequent milk composition. Visualization was performed using standardized path coefficients, with node color representing sample type. Only significant paths are shown in the figure, with blue arrows indicating positive associations and red arrows showing negative associations.

## Data Availability

The raw metagenomic and 16S rRNA gene sequencing data analyzed in this study have been deposited at the NCBI database under BioProject accession code PRJNA1113165 and PRJNA1292286. The milk composition data can be accessed upon request to, and the de-identified personal data can be access upon request to.

## Acknowledgements

This research was funded by grant INV-001734 and OPP1175213 from Bill & Melinda Gates Foundation. Lishi Deng is supported by the China Scholarship Council (Grant No. 202207650056). The funders had no role in the design and conduct of the study; in the collection, management, analysis, and interpretation of the data; or in the preparation, review, or approval of the manuscript. The authors thank all participants from Burkina Faso and the data collection team.

## Author contributions

L.D. conceived the study with input from all authors. L.D. designed and implemented the computational methodology. T.C.D. supervised the study. T.C.D., L.C.T., and C.L. acquired funding and oversaw the implementation of the MISAME-III trial. M.B.A. supervised human milk sample analysis and coordinated data access. J.L.S. supervised stool sample shotgun metagenomic sequencing. K.F. and M.B.M. generated and preprocessed the milk 16S rRNA gene sequencing data. L.H.A. and D.H. generated data on human milk minerals and vitamins. L.B., B.R., and C.Y. generated data on human milk oligosaccharides and bioactive components. T.C.D. and A.M. supported computational methodology. T.C.D., B.D.M., C.L., and M.B.A. contributed to result interpretation. L.D. drafted the manuscript with input from all authors.

