## Supplementary Information for "Time-specific bidirectional links between maternal microbiome, milk composition and infant gut microbiota"

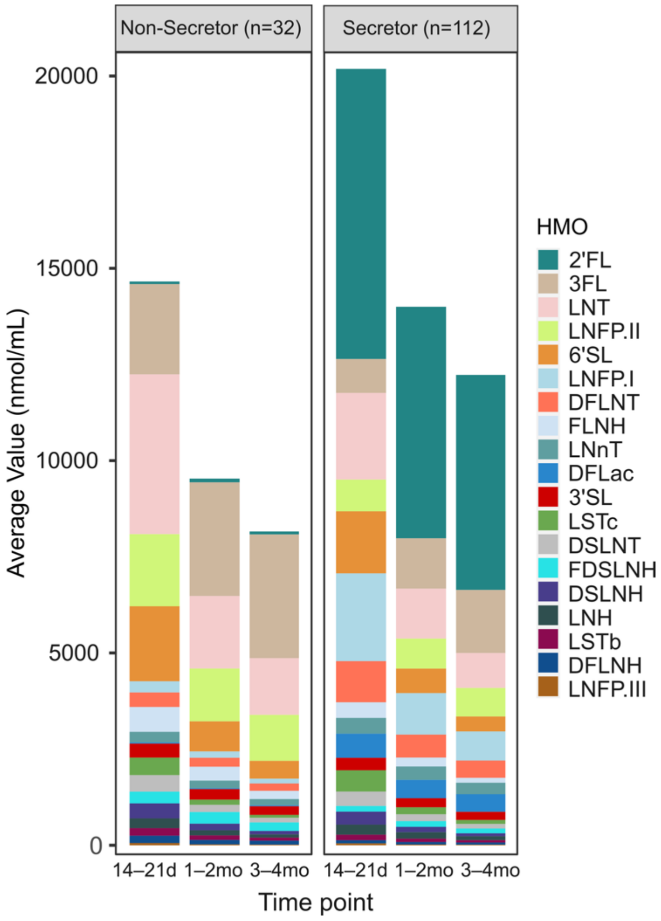


**Figure S1. Stacked-bar plots of mean human milk oligosaccharide concentrations over early lactation, stratified by maternal secretor phenotype.** Mean concentrations of individual HMOs (nmol/mL) were measured in breast milk collected at 14–21 days (14–21 d), 1–2 months (1–2 mo) and 3–4 months (3–4 mo) postpartum. Non-secretor (left panel; n = 32) and secretor (right panel; n = 112) mothers are shown separately. Bars at each timepoint are stacked to display the contribution of each HMO species (color-coded as in the inset key) to total HMO abundance (total bar height).

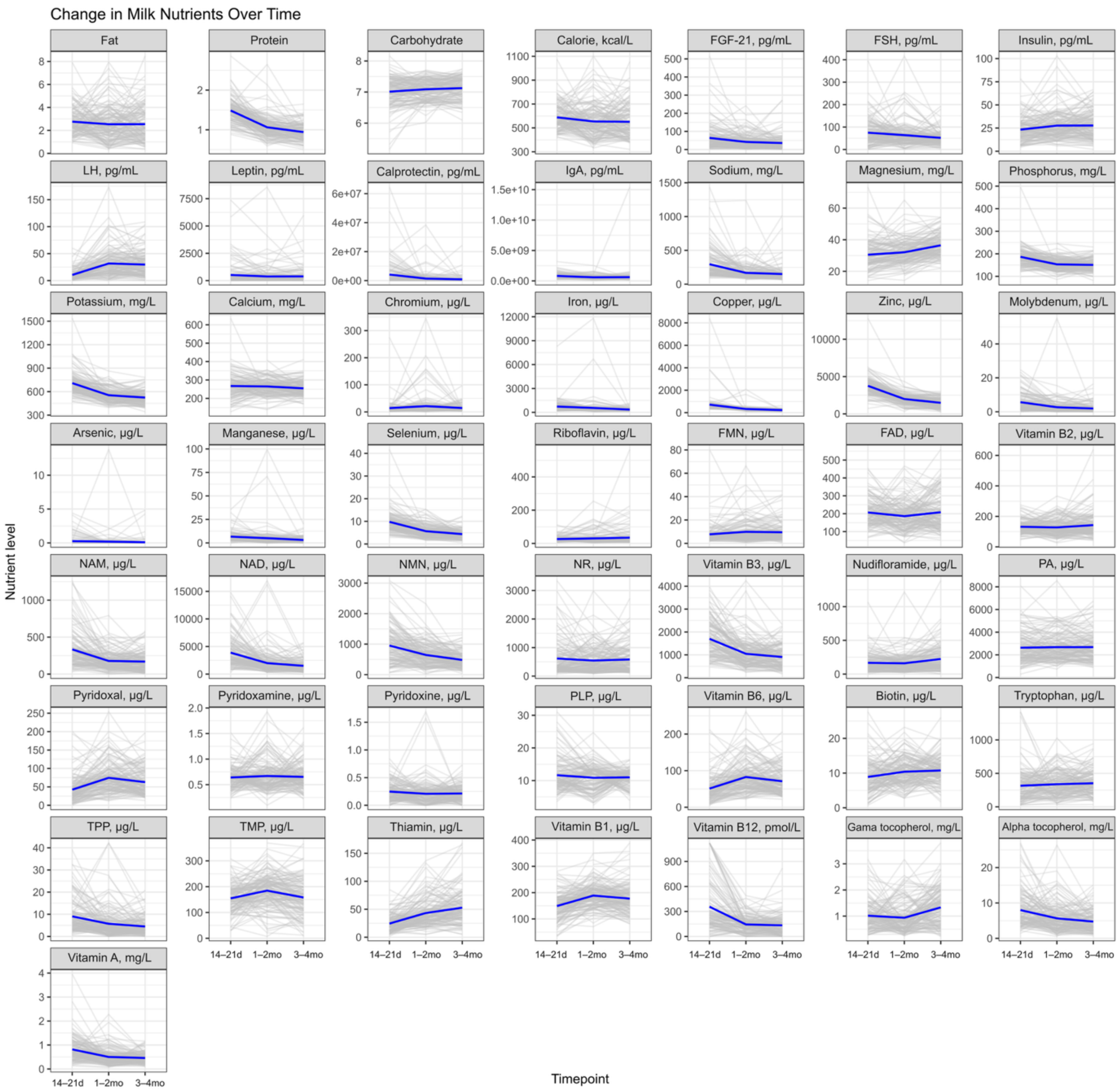


**Figure S2. Longitudinal profiles of breast-milk macronutrients, bioactive proteins, minerals and vitamins over the first four months of lactation**. Milk was sampled from each mother–infant dyad at 14–21 days, 1–2 months and 3–4 months postpartum. In each panel, individual trajectories (one line per dyad) are shown in light grey and the group mean at each timepoint is overlaid in solid blue. Panels are arranged by nutrient class (macronutrients, bioactive proteins, minerals, vitamins), and each y-axis denotes the concentration units for that analyte (g/L for macronutrients; mg/L for proteins and vitamins; µg/L for minerals).

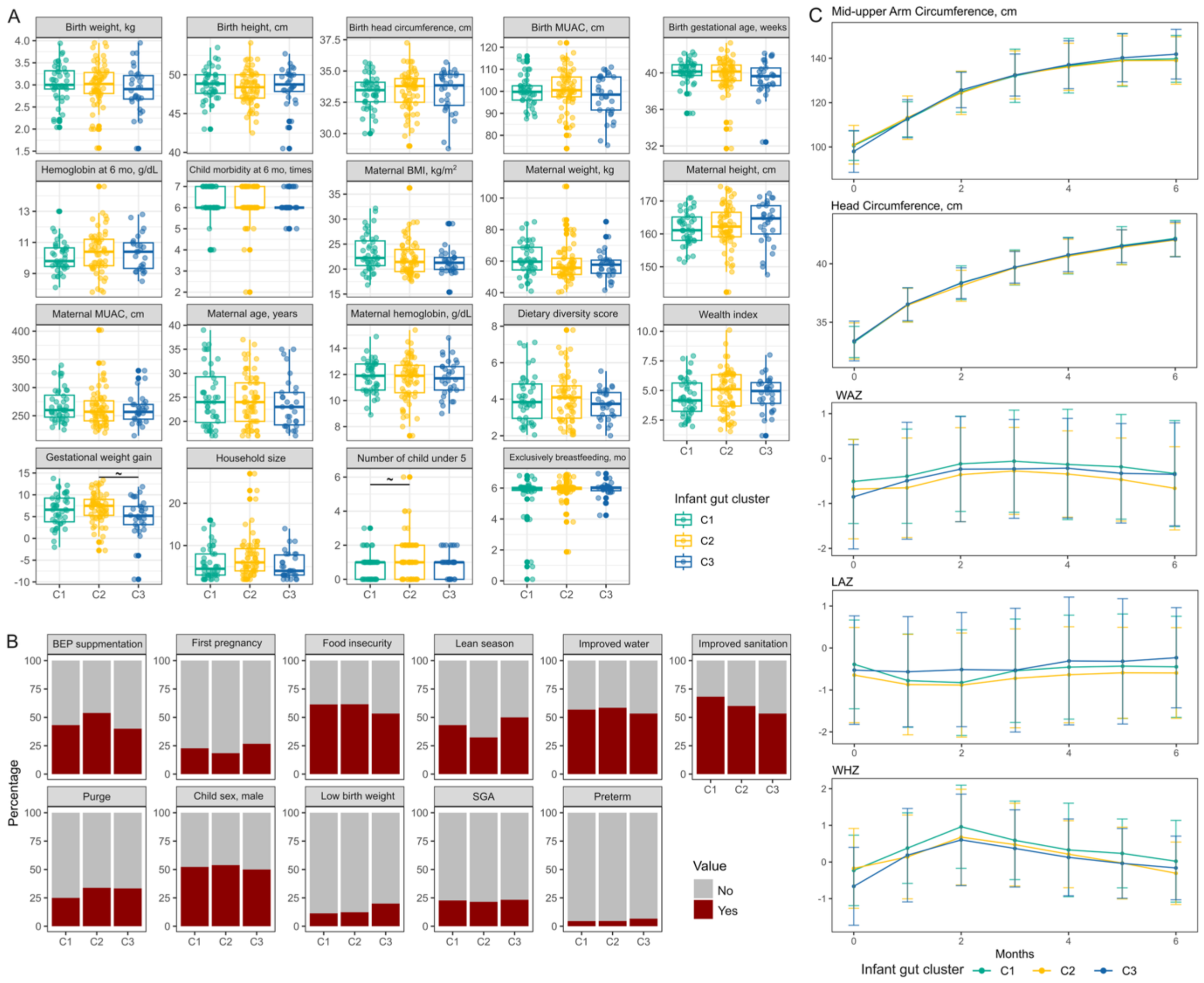


**Figure S3. Maternal and infant baseline characteristics and early growth trajectories by infant gut-microbiome cluster.** Breast-milk samples and clinical data were collected from mother–infant dyads and infants were assigned to one of three gut-microbiome clusters (C1–C3).

**(A)** Continuous characteristics for infants [birth weight, height, head circumference, mid-upper arm circumference (MUAC), gestational age, hemoglobin levels, and morbidity at 6 months], and mothers (body mass index, weight, height, MUAC, age, hemoglobin levels, gestational weight gain, and exclusively breastfeeding duration), plus household and environmental metrics (dietary diversity score, wealth index, household size, number of children under 5 years old in the house). Boxplots show the median (central line), 25$^{th}$ and 75$^{th}$ percentile (box) and 1.5 × interquartile range (IQR) (whiskers). "~" indicates unadjusted $p < 0.05$ before multiple testing correction (Benjamini–Hochberg).

**(B)** Proportion of infants in each cluster with selected binary traits: maternal balanced energy-protein (BEP) supplementation, first pregnancy, food insecurity, born at lean season, improved water and sanitation, purge practice (a traditional custom in rural Burkina Faso where infants undergo a rectal douche to cleanse their bowels), child sex, low birth weight (<2.5kg), small for gestational age (SGA), and preterm birth. Bars represent the percentage of participants with each trait in each cluster.

**(C)** Longitudinal infant growth trajectories from birth to 6 months for MUAC, head circumference, weight-for-age z-score (WAZ), length-for-age z-score (LAZ), and weight-for-length z-score (WHZ), stratified by infant cluster. Lines show the cluster mean at each age; error bars represent standard errors.

Clusters are color-coded as indicated. P-values comparing C1–C3 (panels A and B) were adjusted by the Benjamini–Hochberg method.

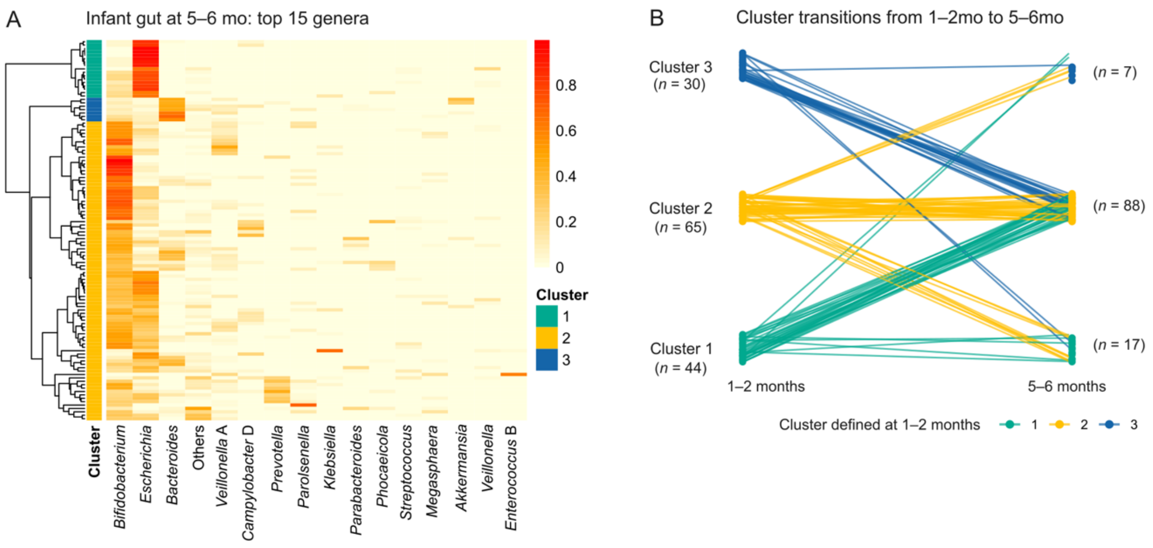


**Figure S4. Characterization and temporal transitions of infant gut microbiome clusters**

(A) Hierarchical clustering heatmap (Bray-Curtis distance, Ward's method) of the top 15 genera in infant stool samples at 5–6 months, identifying three microbial community types. Low-abundance genera were grouped as "Other".

(B) Alluvial plot illustrating transitions in cluster membership from 1–2 months to 5–6 months. Each line represents an individual infant, colored by their cluster identity at 1–2 months.

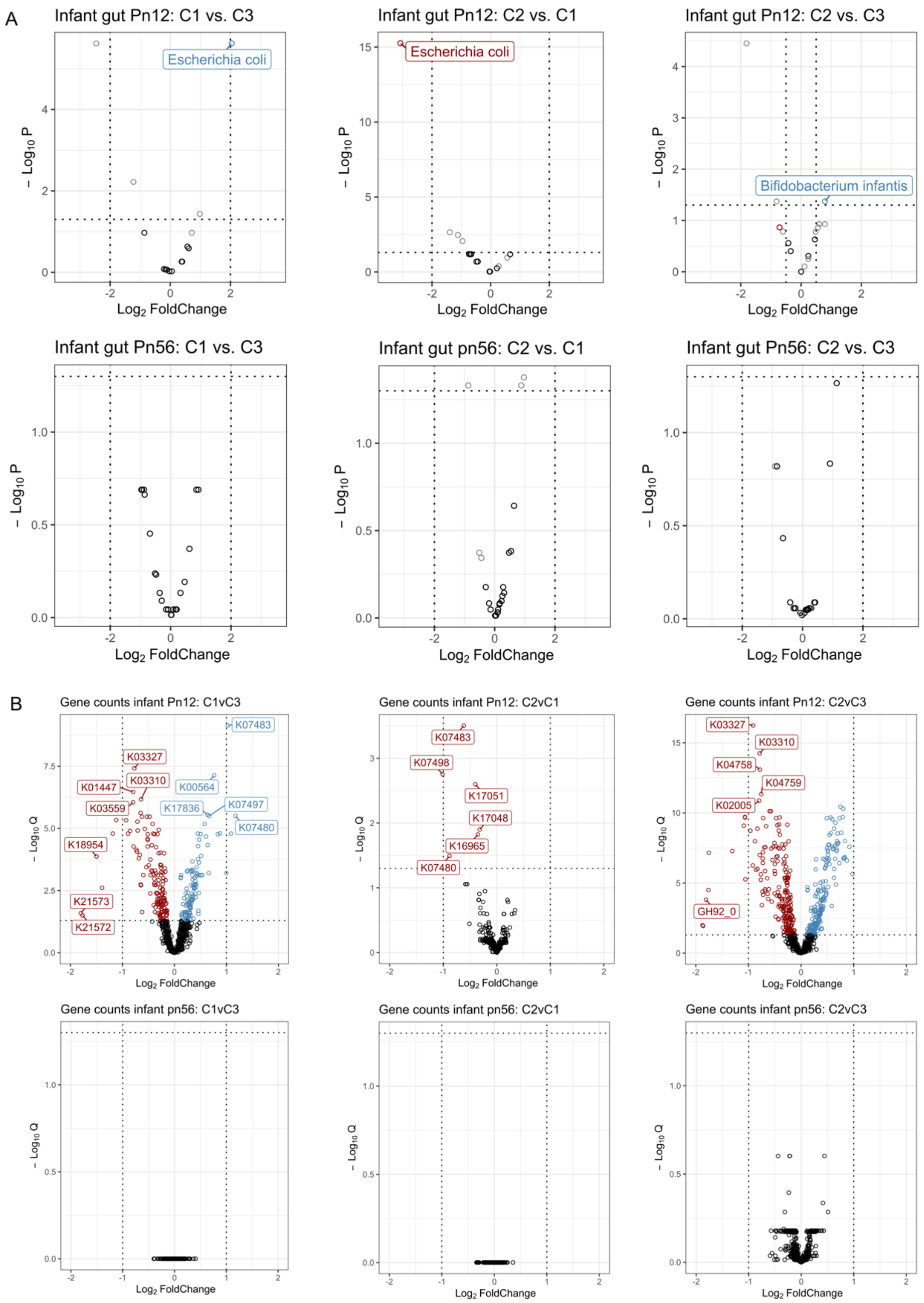


**Figure S5. Volcano plots summaries of differential abundance for infant gut microbial species and functional genes across infant gut clusters (defined at 1–2 months).**

**(A)** Differential abundance of species in the infant gut microbiome across clusters at 1–2 months and 5–6 months, assessed using ANCOM-BC2. Volcano plots display $\log_2$ fold changes versus $-\log_{10}$(p-values) for each pairwise comparison. Selected species of interest are annotated.

**(B)** Differential abundance of functional gene counts (KEGG orthologs) across infant clusters at 1–2 months and 5–6 months. Volcano plots show $\log_2$ fold changes versus $-\log_{10}$(adjusted q-values) from pairwise comparisons. Only gene features with adjusted q-value < 0.05 are highlighted.

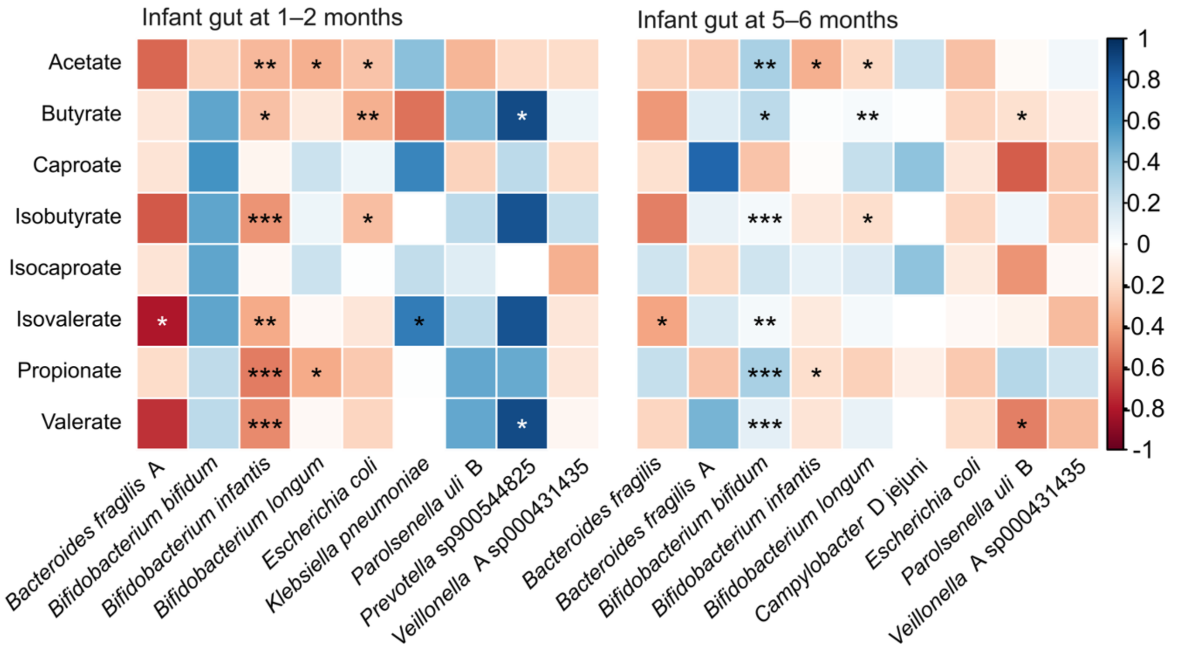


**Figure S6. Correlation heatmap between fecal short chain fatty acids and dominant infant gut species.** Heatmaps of Spearman's rank correlation coefficients between concentrations of eight short-chain fatty acids (SCFAs; rows) and the relative abundances of gut bacterial species with mean abundance >5% (columns) in infant stool at 1–2 months (left) and 5–6 months (right). Tile color corresponds to the correlation coefficient. ***p(FDR) < 0.001; **p(FDR) < 0.01; *p(FDR) <0.05; ~ p(unadjusted) < 0.05.

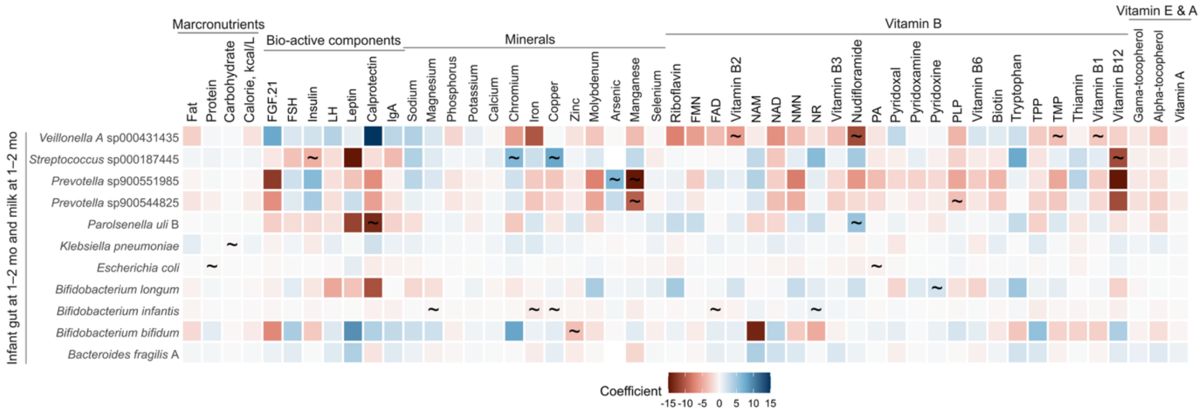


**Figure S7. Heatmap of associations between milk nutrients and infant gut microbial species at 1–2 months.** Associations between concentrations of individual milk nutrients at 1–2 months postpartum and the relative abundances of dominant infant-gut bacterial species (>5 % mean abundance) at 1–2 months were estimated by linear regression, with all models adjusted for maternal balanced energy–protein (BEP) supplementation. Each tile shows the regression coefficient (β), with red indicating a positive association and blue indicating a negative association; tile intensity reflects the magnitude of β. "~" indicates unadjusted $p < 0.05$ before multiple testing correction (Benjamini–Hochberg).

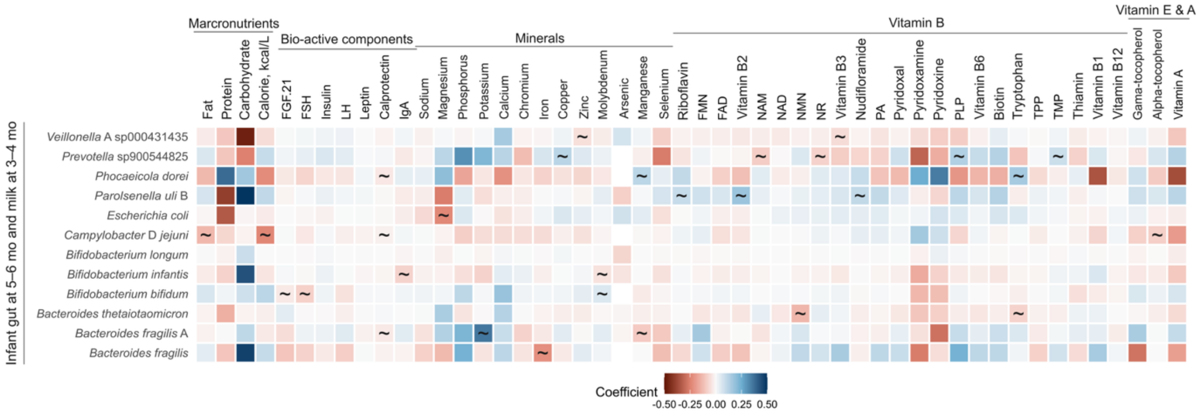


**Figure S8. Heatmap of associations between 3–4-month milk nutrients and 5–6-month infant gut microbial species.** Associations between concentrations of individual milk nutrients at 3–4 months postpartum and the relative abundances of dominant infant-gut bacterial species (>5 % mean abundance) at 5–6 months were estimated by linear regression, with all models adjusted for maternal balanced energy–protein (BEP) supplementation. Each tile shows the regression coefficient (β), with red indicating a positive association and blue indicating a negative association; tile intensity reflects the magnitude of β. "~" indicates unadjusted $p$ < 0.05 before multiple testing correction (Benjamini–Hochberg).

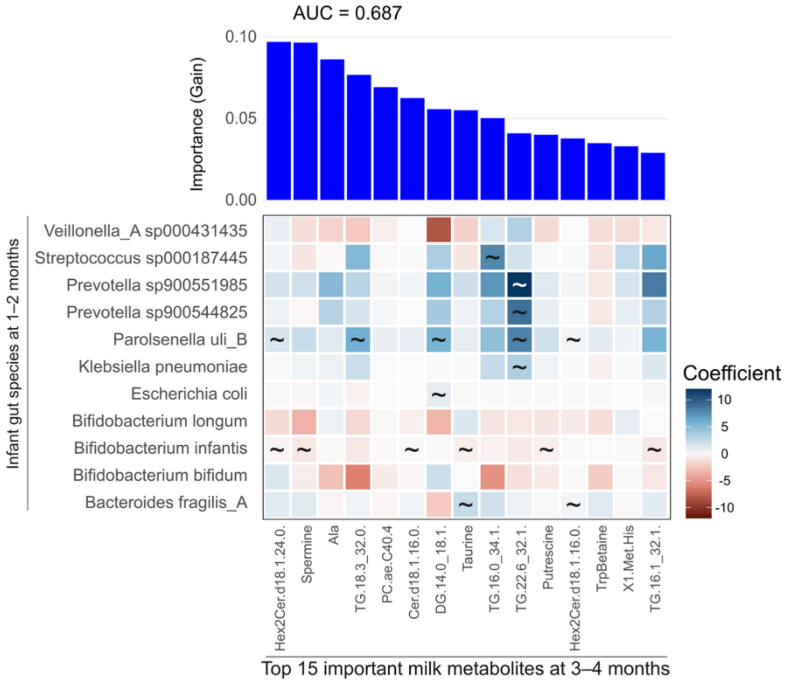


**Figure S9. Top 15 milk metabolites predictive of infant gut clusters and their associations with infant gut species.** Upper panel: Top 15 most important milk metabolites at 3–4 months postpartum identified by an XGBoost classifier trained to predict infant gut clusters at 1–2 months. Feature importance is based on model gain, with an area under the curve (AUC) of 0.687.

Lower panel: Associations between concentrations of the top 15 milk metabolites at 3–4 months and the relative abundances of infant gut species at 1–2 months. Associations were assessed using linear models adjusting for BEP supplementation. Only infant species with mean relative abundance >5% were included. Color scale represents regression coefficients. "~" indicates unadjusted $p$ < 0.05 before multiple testing correction (Benjamini–Hochberg).

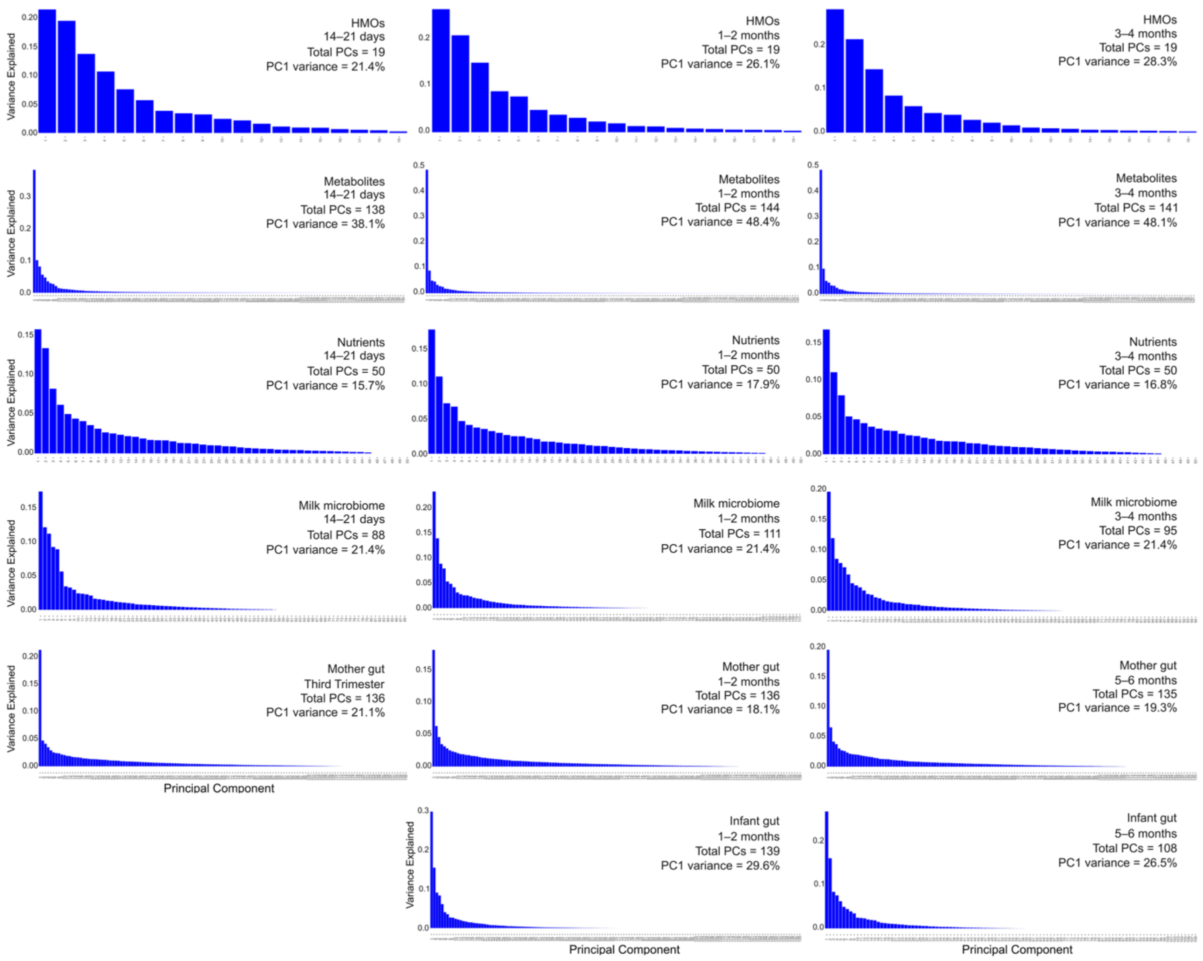


**Figure S10. Distributions of first ordination–axis scores for milk nutrient and microbiome datasets across sampling timepoints.** Histograms display the distribution of individual-sample scores along the first ordination axis (PC1) for each dataset at its respective collection time. For the milk omics panels (rows 1–4: HMO, lipidomics, nutrients and metabolites), samples collected at 14–21 days, 1–2 months and 3–4 months postpartum were analyzed by PCA on standardized concentration data; the percentage of variance captured by PC1 is noted in each panel. The milk microbiome (row 5), maternal gut microbiome (row 6) and infant gut microbiome (row 7) panels show PC1 score from PcoA of weighted UniFrac distance, with the percent variance explained indicated.

**Table S1: Baseline characteristics of MISAME-III participants and the subset participants, related to Figure 1**

| Characteristics | MISAME-III | Subset | | | |
|---|---|---|---|---|---|
| | Overall (*n* = 1897) | C1 (*n* = 44) | C2 (*n* = 65) | C3 (*n* = 30) | $P_{FDR}$ value |
| **Health center catchment area, n (%)** | | | | | 0.56 |
| Boni | 433 (22.8) | 6 (13.6) | 11 (16.9) | 5 (16.7) | |
| Dohoun | 200 (10.5) | 3 (6.8) | 6 (9.2) | 6 (20.0) | |
| Dougoumato II | 346 (18.2) | 7 (15.9) | 14 (21.5) | 6 (20.0) | |
| Karaba | 193 (10.2) | 6 (13.6) | 10 (15.4) | 2 (6.7) | |
| Kari | 350 (18.5) | 9 (20.5) | 13 (20.0) | 8 (26.7) | |
| Koumbia | 375 (19.8) | 13 (19.5) | 11 (16.9) | 10 (20.0) | |
| **Household level** | | | | | |
| Household food insecurity*, n (%) | 1277 (67.5) | 27 (61.4) | 40 (61.5) | 16 (53.3) | 0.75 |
| Wealth index, 0 to 10 points | 4.59 ± 1.76 | 4.38 ± 1.57 | 4.89 ± 1.84 | 4.71 ± 1.45 | 0.29 |
| Household size, number | 6.26 ± 4.38 | 5.68 ± 3.77 | 7.09 ± 5.02 | 5.2 ± 3.17 | 0.08 |
| Number of children under five | 1.05 ± 0.98 | 0.82 ± 0.76 | 1.33 ± 1.20 | 0.93 ± 0.74 | 0.02 |
| Improved water, n (%) | 1193 (62.9) | 25 (56.8) | 38 (58.5) | 16 (53.3) | 0.88 |
| Improved sanitation, n (%) | 1135 (59.8) | 30 (68.2) | 39 (60.0) | 16 (53.3) | 0.43 |
| **Mother** | | | | | |
| Ethnic group, n (%) | | | | | 0.42 |
| Bwaba | 1086 (57.3) | 24 (54.5) | 30 (46.2) | 19 (63.3) | |
| Mossi | 662 (34.9) | 13 (29.5) | 26 (40.0) | 10 (33.3) | |
| Others | 148 (7.8) | 7 (15.9) | 9 (13.8) | 1 (3.3) | |
| Religion of pregnant woman, n (%) | | | | | 0.57 |
| Animist | 432 (22.8) | 11 (25.0) | 12 (18.5) | 8 (26.7) | |
| Muslim | 801 (42.2) | 18 (40.9) | 34 (52.3) | 11 (36.7) | |
| Catholic | 261 (13.8) | 4 (9.1) | 2 (3.1) | 3 (10.0) | |
| Protestant | 335 (17.7) | 10 (22.7) | 15 (23.1) | 8 (26.7) | |
| No religion, no animist | 68 (3.6) | 1 (2.3) | 2 (3.1) | 0 (0.0) | |
| Primary education and above, n (%) | 797 (42.0) | 18 (40.9) | 26 (40.0) | 13 (43.3) | 0.73 |
| Number of jobs, % | | | | | 0.78 |
| 0 | 1152 (60.7) | 23 (52.3) | 40 (61.5) | 18 (60.0) | |
| 1 | 671 (35.4) | 19 (43.2) | 24 (36.9) | 11 (36.7) | |
| 2 and above | 74 (3.9) | 2 (4.5) | 1 (1.5) | 1 (3.3) | |
| Parity, % | | | | | 0.66 |
| 0 | 429 (22.6) | 10 (22.7) | 12 (18.5) | 8 (26.7) | |
| 1-2 | 654 (34.5) | 14 (31.8) | 29 (44.6) | 11 (36.7) | |
| 3 or more | 814 (42.9) | 20 (45.5) | 24 (36.9) | 11 (36.7) | |
| BEP supplementation, n (%) | | | | | 0.37 |
| Prenatal intervention | 462 (24.4) | 0 (0.0) | 0 (0.0) | 0 (0.0) | |
| Postnatal intervention | 471 (24.8) | 0 (0.0) | 0 (0.0) | 0 (0.0) | |
| Pre- and postnatal intervention | 475 (25.0) | 19 (43.2) | 35 (53.8) | 12 (40.0) | |
| Control | 489 (25.8) | 25 (56.8) | 30 (46.2) | 18 (60.0) | |
| Age, years | 25.0 ± 6.17 | 25.07 ± 6.15 | 24.31 ± 5.18 | 23.73 ± 5.32 | 0.58 |
| Weight, kg | 58.17 ± 8.67 | 60.90 ± 10.43 | 58.48 ± 11.45 | 57.66 ± 8.88 | 0.36 |
| Height, cm | 162.54 ± 6.03 | 161.72 ± 5.03 | 162.28 ± 6.42 | 163.31 ± 6.60 | 0.54 |
| Body mass index, kg/m$^2$ | 21.99 ± 2.87 | 23.25 ± 3.68 | 22.12 ± 3.49 | 21.57 ± 2.72 | 0.09 |
| Mid-upper arm circumference, cm | 262.02 ± 26.63 | 268.30 ± 28.71 | 263.45 ± 32.09 | 260.67 ± 24.86 | 0.52 |
| Hemoglobin, g/dL | 11.34 ± 1.51 | 11.84 ± 1.23 | 11.66 ± 1.63 | 11.73 ± 1.40 | 0.81 |
| Anemia, hemoglobin <11 g/dL, n (%) | 734 (38.7) | 15 (34.1) | 25 (38.5) | 10 (33.3) | 0.85 |
| Gestational weight gain, kg | 6.17 ± 3.52 | 6.43 ± 3.63 | 7.03 ± 3.28 | 4.96 ± 4.44 | 0.04 |
| Dietary diversity score, 0 to 10 points | 3.96 ± 1.61 | 4.06 ± 1.31 | 4.06 ± 1.32 | 3.71 ± 0.83 | 0.39 |
| **Infant** | | | | | |
| Sex, male, n (%) | 862 (49.6) | 23 (52.3) | 35 (53.8) | 15 (50.0) | 0.97 |
| Born in lean season, n (%) | 527 (31.8) | 19 (43.2) | 21 (32.3) | 15 (50.0) | 0.21 |
| Birth weight, kg | 3.01 ± 0.44 | 3.05 ± 0.42 | 3.00 ± 0.49 | 2.90 ± 0.51 | 0.42 |
| Birth length, cm | 48.29 ± 2.19 | 48.70 ± 2.01 | 48.32 ± 2.21 | 48.47 ± 2.55 | 0.68 |
| Birth head circumference, cm | 3.42 ± 1.58 | 33.29 ± 1.35 | 33.42 ± 1.57 | 33.36 ± 1.73 | 0.91 |
| Birth mid-upper arm circumference, cm | 100.76 ± 8.29 | 100.31 ± 6.75 | 101.08 ± 9.16 | 97.65 ± 9.81 | 0.2 |
| Gestational age at birth, weeks | 39.91 ± 1.75 | 40.10 ± 1.41 | 39.79 ± 1.88 | 39.47 ± 1.92 | 0.32 |
| Hemoglobin level at 6 months, g/dL | 10.41 ± 1.36 | 10.06 ± 0.99 | 10.38 ± 1.39 | 10.34 ± 1.19 | 0.42 |
| Morbidity at 6 months, times | 6.40 ± 1.77 | 6.27 ± 0.82 | 6.18 ± 0.85 | 6.13 ± 0.57 | 0.73 |

Data are presented as n (%) for categorical variables and mean ± sd for continuous variables.
Comparisons were performed using Fisher's exact test for categorical variables and ANOVA for continuous variables.
BEP: balanced energy-protein.

**Table S2. Redundancy analysis (RDA) of the infant gut microbiome at 1–2 months including milk nutrients at 14–21 days, related to Figure 7**

| Feature abbreviation | Full name | Variance explained | F-value | p-value |
|---|---|---|---|---|
| FAT | Total fat | 0.091 | 0.563 | 0.568 |
| PROTEIN | Total protein | 0.342 | 2.123 | 0.113 |
| CARBOHYDRATE | Total carbohydrate | 0.096 | 0.596 | 0.533 |
| FGF.21 | Fibroblast growth factor 21 | 0.011 | 0.066 | 0.997 |
| FSH | Follicle-stimulating hormone | 0.058 | 0.358 | 0.731 |
| Insulin | Insulin | 0.082 | 0.512 | 0.619 |
| LH | Luteinizing hormone | 0.041 | 0.254 | 0.841 |
| Leptin | Leptin | 0.342 | 2.127 | 0.124 |
| Calprotectin | Milk calprotectin | 0.235 | 1.464 | 0.216 |
| IgA | Immunoglobulin A | 0.343 | 2.131 | 0.102 |
| Na | Sodium | 0.114 | 0.706 | 0.502 |
| Mg | Magnesium | 0.194 | 1.204 | 0.278 |
| P. | Phosphorus | 0.057 | 0.351 | 0.758 |
| K. | Potassium | 0.074 | 0.462 | 0.675 |
| Ca | Calcium | 0.037 | 0.231 | 0.888 |
| Cr | Chromium | 0.036 | 0.222 | 0.872 |
| Fe | Iron | 0.334 | 2.078 | 0.110 |
| Cu | Copper | 0.119 | 0.740 | 0.485 |
| Zn | Zinc | 0.049 | 0.307 | 0.807 |
| Mo | Molybdenum | 0.166 | 1.033 | 0.351 |
| As | Arsenic | 0.185 | 1.152 | 0.307 |
| Mn | Manganese | 0.020 | 0.124 | 0.953 |
| Se | Selenium | 0.049 | 0.303 | 0.820 |
| Ribo | Riboflavin | 0.107 | 0.668 | 0.512 |
| FMN | Flavin mononucleotide | 0.026 | 0.162 | 0.926 |
| FAD | Flavin adenine dinucleotide | 0.430 | 2.674 | 0.061 |
| NAM | Nicotinamide | 0.104 | 0.647 | 0.514 |
| NAD | Nicotinamide adenine dinucleotide | 0.155 | 0.966 | 0.367 |
| NMN | Nicotinamide mononucleotide | 0.044 | 0.276 | 0.822 |
| NR | Nicotinamide riboside | 0.129 | 0.804 | 0.436 |
| B3 | Vitamin B3 | 0.066 | 0.408 | 0.704 |
| Nufa | Nudifloramide (niacin catabolite) | 0.281 | 1.748 | 0.166 |
| PA | Pantothenic acid | 0.179 | 1.114 | 0.321 |
| PL | Pyridoxal | 0.100 | 0.620 | 0.540 |
| PM | Pyridoxamine | 0.058 | 0.362 | 0.746 |
| PN | Pyridoxine | 0.087 | 0.541 | 0.605 |
| PLP | Pyridoxal 5-phosphate | 0.301 | 1.874 | 0.147 |
| Bio | Biotin | 0.135 | 0.838 | 0.414 |
| TRP | Tryptophan | 0.051 | 0.315 | 0.800 |
| TPP | Thiamine pyrophosphate | 0.065 | 0.403 | 0.712 |
| TMP | Thiamine monophosphate | 0.046 | 0.284 | 0.827 |
| T | Free thiamin | 0.070 | 0.437 | 0.700 |
| B12 | Vitamin B12 | 0.034 | 0.213 | 0.908 |
| g.tocopherol | Gama-tocopherol (Vitamin E) | 0.073 | 0.455 | 0.658 |
| a.tocopherol | Alpha-tocopherol (Vitamin E) | 0.148 | 0.918 | 0.391 |
| vitamin.A | Vitamin A | 0.268 | 1.664 | 0.174 |

**Table S3. Redundancy analysis (RDA) of the infant gut microbiome at 1–2 months including milk nutrients at 1–2 months, related to Figure 7**

| Feature abbreviation | Full name | Variance explained | F-value | p-value |
|---|---|---|---|---|
| B3 | Vitamin B3 | 0.706 | 4.091 | 0.016 |
| PROTEIN | Total protein | 0.718 | 4.159 | 0.018 |
| TRP | Tryptophan | 0.595 | 3.445 | 0.028 |
| Mg | Magnesium | 0.694 | 4.019 | 0.030 |
| g.tocopherol | Gama-tocopherol (Vitamin E) | 0.609 | 3.527 | 0.035 |
| FAT | Total fat | 0.601 | 3.481 | 0.037 |
| PA | Pantothenic acid | 0.441 | 2.557 | 0.077 |
| Nufa | Nudifloramide (niacin catabolite) | 0.421 | 2.441 | 0.085 |
| Zn | Zinc | 0.294 | 1.706 | 0.168 |
| PLP | Pyridoxal 5-phosphate | 0.296 | 1.717 | 0.178 |
| IgA | Immunoglobulin A | 0.268 | 1.554 | 0.198 |
| a.tocopherol | Alpha-tocopherol (Vitamin E) | 0.250 | 1.446 | 0.223 |
| Calprotectin | Milk calprotectin | 0.221 | 1.278 | 0.239 |
| T | Free thiamin | 0.193 | 1.121 | 0.299 |
| PN | Pyridoxine | 0.184 | 1.068 | 0.311 |
| K. | Potassium | 0.186 | 1.077 | 0.327 |
| Leptin | Leptin | 0.157 | 0.908 | 0.373 |
| Mn | Manganese | 0.155 | 0.900 | 0.400 |
| Fe | Iron | 0.137 | 0.795 | 0.431 |
| NAM | Nicotinamide | 0.148 | 0.856 | 0.438 |
| Na | Sodium | 0.136 | 0.788 | 0.466 |
| Mo | Molybdenum | 0.124 | 0.721 | 0.510 |
| Bio | Biotin | 0.110 | 0.636 | 0.542 |
| vitamin.A | Vitamin A | 0.112 | 0.650 | 0.549 |
| Insulin | Insulin | 0.113 | 0.654 | 0.553 |
| Cu | Copper | 0.095 | 0.552 | 0.589 |
| NR | Nicotinamide riboside | 0.092 | 0.530 | 0.601 |
| Ca | Calcium | 0.090 | 0.523 | 0.632 |
| NMN | Nicotinamide mononucleotide | 0.078 | 0.452 | 0.694 |
| P. | Phosphorus | 0.071 | 0.411 | 0.696 |
| FAD | Flavin adenine dinucleotide | 0.067 | 0.389 | 0.717 |
| As | Arsenic | 0.065 | 0.374 | 0.728 |
| Se | Selenium | 0.057 | 0.328 | 0.768 |
| TMP | Thiamine monophosphate | 0.058 | 0.333 | 0.778 |
| FGF.21 | Fibroblast growth factor 21 | 0.056 | 0.324 | 0.784 |
| FSH | Follicle-stimulating hormone | 0.047 | 0.273 | 0.810 |
| CARBOHYDRATE | Total carbohydrate | 0.052 | 0.304 | 0.818 |
| PL | Pyridoxal | 0.045 | 0.259 | 0.839 |
| LH | Luteinizing hormone | 0.045 | 0.261 | 0.842 |
| Cr | Chromium | 0.032 | 0.186 | 0.879 |
| FMN | Flavin mononucleotide | 0.038 | 0.220 | 0.881 |
| TPP | Thiamine pyrophosphate | 0.032 | 0.187 | 0.914 |
| B12 | Vitamin B12 | 0.031 | 0.180 | 0.922 |
| PM | Pyridoxamine | 0.028 | 0.161 | 0.939 |
| Ribo | Riboflavin | 0.020 | 0.119 | 0.970 |
| NAD | Nicotinamide adenine dinucleotide | 0.012 | 0.068 | 0.999 |

**Table S4. Redundancy analysis (RDA) of the infant gut microbiome at 1–2 months including all maternal, milk and infant factors, related to Figure 7**

| Feature abbreviation | Full name | Variance explained | F-value | p-value |
|---|---|---|---|---|
| D1421_DFLac | Difucosyllactose at 14–21 days | 1.090 | 7.549 | 0.002 |
| D1421_X3.SL | 3’-sialyllactose at 14–21 days | 0.747 | 5.170 | 0.005 |
| Pn12_DFLac | Difucosyllactose at 1–2 months | 0.597 | 4.134 | 0.021 |
| D1421_DSLNT | Disialyllactose-*N*-tetraose at 14–21 days | 0.557 | 3.855 | 0.022 |
| Milk_Veillonella_A | Milk genus *Veillonella* A | 0.540 | 3.742 | 0.026 |
| Pn12_LNT | Lacto-*N*-tetrose at 1–2 months | 0.510 | 3.534 | 0.028 |
| Tri3_C941 sp004557565 | Maternal gut species C941 sp004557565 at third trimester | 0.511 | 3.540 | 0.03 |
| Milk_Shannon | Milk Shannon diversity | 0.452 | 3.133 | 0.036 |
| Milk_Erysipelatoclostridium | Milk genus *Erysipelatoclostridium* | 0.463 | 3.205 | 0.044 |
| M_Height | Maternal height at inclusion | 0.424 | 2.935 | 0.047 |
| D1421_Fuc | HMO bound fucose at 14–21 days | 0.454 | 3.141 | 0.05 |
| Tri3_Prevotella sp900548535 | Maternal gut species *Prevotella* sp900548535 at third trimester | 0.433 | 3.000 | 0.062 |
| M_GestationalWeightGain | Maternal gestational weight gain | 0.403 | 2.789 | 0.063 |
| M_WealthIndex | Maternal wealth index (1 to 10) | 0.384 | 2.657 | 0.067 |
| D1421_FDSLNH | Fucodisialyllacto-*N*-hexaose at 14–21 days | 0.351 | 2.431 | 0.078 |
| Tri3_Prevotella sp900556795 | Maternal gut species *Prevotella* sp900556795at third trimester | 0.330 | 2.287 | 0.086 |
| Pn12_Diversity | HMO diversity at 1–2 months | 0.338 | 2.340 | 0.1 |
| D1421_LNH | Lacto-*N*-hexaose at 14–21 days | 0.311 | 2.154 | 0.113 |
| D1421_X2.FL | 2’-fucosyllactose at 14–21 days | 0.267 | 1.852 | 0.154 |
| Tri3_RC9 sp000433355 | Maternal gut species RC9 sp000433355 at third trimester | 0.278 | 1.926 | 0.156 |
| Pn12_DFLNT | Difucosyllacto-*N*-tetraose at 1–2 months | 0.260 | 1.798 | 0.159 |
| D1421_LNFP.I | Lacto-*N*-fucopentaose I at 14–21 days | 0.259 | 1.791 | 0.162 |
| Pn12_X3.SL | 3’-Sialyllactose at 1–2 months | 0.223 | 1.544 | 0.195 |
| Pn12_LSTc | Sialyllactose-*N*-tetraose c at 1–2 months | 0.233 | 1.616 | 0.199 |
| SGA | Small for gestational age | 0.209 | 1.446 | 0.227 |
| C_GestationalAge | Infant gestational age at birth | 0.212 | 1.466 | 0.243 |
| Pn12_FLNH | Fucosyllacto-*N*-hexaose at 1–2 months | 0.211 | 1.461 | 0.243 |
| C_Weight | Infant weight at birth | 0.204 | 1.416 | 0.254 |
| Pn12_LNFP.II | Lacto-*N*-fucopentaose II at 1–2 months | 0.204 | 1.414 | 0.255 |
| M_MUAC | Maternal mid-upper arm circumference at inclusion | 0.208 | 1.439 | 0.265 |
| Tri3_Prevotella sp000434515 | Maternal gut species *Prevotella* sp000434515 at third trimester | 0.189 | 1.311 | 0.272 |
| D1421_LNnT | Lacto-*N*-neotetraose at 14–21 days | 0.187 | 1.296 | 0.282 |
| Pn12_FDSLNH | Fucodisialyllacto-N-hexaose at 1–2 months | 0.170 | 1.174 | 0.286 |
| D1421_SUM | Total HMO concentration at 14–21 days | 0.182 | 1.262 | 0.298 |
| HH_Size | Household size | 0.172 | 1.192 | 0.302 |
| D1421_LNFP.II | Lacto-*N*-fucopentaose II at 14–21 days | 0.168 | 1.160 | 0.305 |
| Milk_Streptococcus | Milk genus *Streptococcus* | 0.173 | 1.199 | 0.314 |
| Milk_Parolsenella | Milk genus *Parolsenella* | 0.160 | 1.107 | 0.328 |
| Milk_Collinsella | Milk genus *Collinsella* | 0.166 | 1.148 | 0.331 |
| Tri3_Observed | Maternal gut observed richness at third trimester | 0.162 | 1.119 | 0.351 |
| C_HeadCircumference | Infant head circumference at birth | 0.148 | 1.024 | 0.353 |
| M_Weight | Maternal weight at inclusion | 0.157 | 1.085 | 0.357 |
| Pn12_LNnT | Lacto-*N*-neotetraose at 1–2 months | 0.145 | 1.006 | 0.359 |
| D1421_X3FL | 3-Fucosyllactose at 14–21 days | 0.145 | 1.005 | 0.363 |
| Tri3_Prevotella sp900551275 | Maternal gut species *Prevotella* sp900551275 at third trimester | 0.138 | 0.956 | 0.384 |
| Pn12_Sia | HMO bound sialic acids at 1–2 months | 0.146 | 1.014 | 0.387 |
| Pn12_Secretor | Maternal secretor status | 0.133 | 0.923 | 0.406 |
| HH_Child5 | Number of children under 5 years in household | 0.130 | 0.902 | 0.417 |
| Milk_Prevotella | Milk genus *Prevotella* | 0.123 | 0.851 | 0.438 |
| Pn12_X6.SL | 6’-Sialyllactose at 1–2 months | 0.119 | 0.826 | 0.465 |
| M_BMI | Maternal body mass index at inclusion | 0.118 | 0.817 | 0.467 |
| M_BEP | Maternal balanced energy–protein supplementation (yes/no) | 0.108 | 0.748 | 0.482 |

| | | | | |
|---|---|---|---|---|
| D1421_LNT | Lacto-*N*-tetrose at 14–21 days | 0.112 | 0.775 | 0.498 |
| D1421_X6.SL | 6'-Sialyllactose at 14–21 days | 0.110 | 0.760 | 0.498 |
| Milk_Lactobacillus | Milk genus *Lactobacillus* | 0.109 | 0.754 | 0.518 |
| Pn12_X2.FL | 2'-Fucosyllactose at 1–2 months | 0.102 | 0.703 | 0.522 |
| Preterm | Preterm birth | 0.109 | 0.753 | 0.523 |
| Tri3_Prevotella sp002299635 | Maternal gut species *Prevotella* sp002299635 at third trimester | 0.101 | 0.698 | 0.531 |
| LBW | Low birth weight | 0.100 | 0.694 | 0.545 |
| C_MUAC | Infant mid-upper arm circumference at birth | 0.096 | 0.666 | 0.548 |
| Pn12_SUM | Total HMO concentration at 1–2 months | 0.100 | 0.696 | 0.55 |
| Milk_Pauljensenia | Milk genus *Pauljensenia* | 0.091 | 0.629 | 0.582 |
| M_Age | Maternal age at inclusion | 0.087 | 0.601 | 0.601 |
| Pn12_X3FL | 3-Fucosyllactose at 1–2 months | 0.082 | 0.571 | 0.601 |
| FirstPregnancy | First pregnancy (yes/no) | 0.091 | 0.627 | 0.609 |
| M_Hemoglobin | Maternal hemoglobin level at inclusion | 0.084 | 0.583 | 0.633 |
| C_Height | Infant length/height at inclusion | 0.077 | 0.533 | 0.662 |
| Milk_Limosilactobacillus | Milk genus *Limosilactobacillus* | 0.066 | 0.456 | 0.678 |
| D1421_Sia | HMO bound sialic acids at 14–21 days | 0.066 | 0.458 | 0.687 |
| Tri3_Prevotella sp000436035 | Maternal gut species *Prevotella* sp000436035 at third trimester | 0.072 | 0.500 | 0.694 |
| D1421_DFLNT | Difucosyllacto-*N*-tetraose at 14–21 days | 0.069 | 0.478 | 0.695 |
| Tri3_Shannon | Maternal gut Shannon diversity at third trimester | 0.068 | 0.470 | 0.7 |
| LeanSeason | Infant born in lean season (yes/no) | 0.071 | 0.492 | 0.711 |
| D1421_LSTc | Sialyllactose-*N*-tetraose c at 14–21 days | 0.071 | 0.490 | 0.715 |
| D1421_DSLNH | Disialyllacto-*N*-hexaose at 14–21 days | 0.070 | 0.482 | 0.723 |
| D1421_FLNH | Fucosyllacto-*N*-hexaose at 14–21 days | 0.062 | 0.432 | 0.733 |
| Pn12_LNFP.I | Lacto-*N*-fucopentaose I at 1–2 months | 0.067 | 0.463 | 0.739 |
| Pn12_Fuc | HMO bound fucose at 1–2 months | 0.060 | 0.417 | 0.746 |
| Tri3_Prevotella copri | Maternal gut species *Prevotella copri* at third trimester | 0.059 | 0.408 | 0.749 |
| D1421_Diversity | HMO diversity at 14–21 days | 0.060 | 0.413 | 0.76 |
| ExclusivelyBFmonths | Duration of exclusive breastfeeding (months) | 0.053 | 0.364 | 0.772 |
| Milk_Rothia | Milk genus *Rothia* | 0.046 | 0.319 | 0.828 |
| Purge | Infant purge practice (yes/no) | 0.045 | 0.314 | 0.85 |
| M_DietaryDiversity | Maternal dietary diversity score | 0.040 | 0.280 | 0.871 |
| Tri3_Unknown_Prevotella | Maternal gut unknown *Prevotella* group at third trimester | 0.038 | 0.264 | 0.878 |
| Pn12_DSLNH | Disialyllacto-*N*-hexaose at 1–2 months | 0.034 | 0.237 | 0.913 |
| Pn12_DSLNT | Disialyllacto-*N*-tetraose at 1–2 months | 0.024 | 0.169 | 0.951 |
| Milk_Observed | Milk observed richness | 0.024 | 0.167 | 0.967 |
| Pn12_LNH | Lacto-*N*-hexaose at 1–2 months | 0.018 | 0.122 | 0.978 |

**Table S5. Redundancy analysis (RDA) of the infant gut microbiome at 5–6 months including all maternal, milk and infant factors, related to Figure 7**

| Feature abbreviation | Full name | Variance explained | F-value | p-value |
|---|---|---|---|---|
| Fe | Iron | 1.311 | 17.504 | 0.001 |
| NAD | Nicotinamide adenine dinucleotide | 0.787 | 10.512 | 0.001 |
| FMN | Flavin mononucleotide | 0.568 | 7.583 | 0.002 |
| FDSLNH | Fucodisialyllacto-*N*-hexaose | 0.482 | 6.438 | 0.007 |
| B3 | Vitamin B3 | 0.445 | 5.938 | 0.008 |
| T | Free thiamine | 0.484 | 6.461 | 0.010 |
| Cu | Copper | 0.296 | 3.959 | 0.023 |
| TG.22.6_32.1. | Triglyceride (22:6/32:1) | 0.276 | 3.688 | 0.027 |
| hh_size | Household size | 0.299 | 3.988 | 0.030 |
| PM | Pyridoxamine | 0.264 | 3.520 | 0.037 |
| Fuc | HMO bound fucose | 0.275 | 3.672 | 0.041 |
| Se | Selenium | 0.259 | 3.457 | 0.044 |
| Ca | Calcium | 0.249 | 3.322 | 0.045 |
| Ala | Alanine | 0.234 | 3.119 | 0.049 |
| Mn | Manganese | 0.237 | 3.167 | 0.053 |
| B12 | Vitamin B12 | 0.246 | 3.286 | 0.054 |
| X2.FL | 2’-Fucosyllactose | 0.239 | 3.186 | 0.054 |
| Hex2Cer.d18.1.16.0. | Hexosylceramide (d18:1/16:0) | 0.241 | 3.217 | 0.056 |
| X3FL | 3-Fucosyllactose | 0.224 | 2.985 | 0.059 |
| IgA | Immunoglobulin A | 0.204 | 2.726 | 0.060 |
| a.tocopherol | Alpha-tocopherol (Vitamin E) | 0.206 | 2.746 | 0.063 |
| SUM | Total HMO concentration | 0.216 | 2.885 | 0.069 |
| P. | Phosphorus | 0.220 | 2.937 | 0.070 |
| Secretor | Secretor status | 0.206 | 2.754 | 0.071 |
| mddw_10_ave | Average maternal dietary diversity score (10-food group) | 0.210 | 2.809 | 0.072 |
| Na | Sodium | 0.199 | 2.653 | 0.076 |
| Ribo | Riboflavin | 0.185 | 2.474 | 0.079 |
| iycf_ebf_age | Infant age at end of exclusive breastfeeding | 0.185 | 2.468 | 0.087 |
| CARBOHYDRATE | Total carbohydrate | 0.200 | 2.676 | 0.093 |
| Nufa | Nudifloramide (niacin catabolite) | 0.183 | 2.441 | 0.098 |
| Mo | Molybdenum | 0.178 | 2.383 | 0.100 |
| X3.SL | 3’-Sialyllactose | 0.168 | 2.247 | 0.119 |
| Spermine | Spermine | 0.180 | 2.400 | 0.123 |
| TG.16.1_32.1. | Triglyceride (16:1/32:1) | 0.157 | 2.093 | 0.125 |
| Gabirthweeks | Gestational age at birth (weeks) | 0.168 | 2.244 | 0.129 |
| FAT | Total fat | 0.150 | 2.001 | 0.136 |
| Diversity | HMO diversity | 0.161 | 2.157 | 0.139 |
| Mg | Magnesium | 0.154 | 2.054 | 0.144 |
| Bio | Biotin | 0.145 | 1.932 | 0.151 |
| Putrescine | Putrescine | 0.143 | 1.914 | 0.156 |
| Cr | Chromium | 0.140 | 1.864 | 0.159 |
| As | Arsenic | 0.153 | 2.044 | 0.159 |
| DSLNT | Disialyllacto-*N*-tetraose | 0.135 | 1.801 | 0.167 |
| FAD | Flavin adenine dinucleotide | 0.139 | 1.851 | 0.169 |
| LSTc | Sialyllactose-*N*-tetraose c | 0.134 | 1.795 | 0.173 |
| HH_food_insecurity | Household food insecurity (yes/no) | 0.133 | 1.779 | 0.184 |
| Calprotectin | Milk calprotectin | 0.137 | 1.829 | 0.185 |
| PA | Pantothenic acid | 0.126 | 1.684 | 0.186 |
| TMP | Thiamine monophosphate | 0.124 | 1.653 | 0.187 |
| hh_child5 | Number of children <5 years in household | 0.119 | 1.587 | 0.205 |
| Taurine | Taurine | 0.121 | 1.622 | 0.208 |
| X6.SL | 6’-Sialyllactose | 0.122 | 1.626 | 0.213 |
| TG.16.0_34.1. | Triglyceride (16:0/34:1) | 0.121 | 1.619 | 0.214 |
| m_hbincl | Maternal hemoglobin at inclusion | 0.112 | 1.491 | 0.222 |
| NMN | Nicotinamide mononucleotide | 0.121 | 1.615 | 0.226 |
| Gravidity | Number of pregnancies | 0.114 | 1.519 | 0.227 |
| NR | Nicotinamide riboside | 0.117 | 1.559 | 0.227 |

| | | | | |
|---|---|---|---|---|
| FSH | Follicle-stimulating hormone | 0.117 | 1.557 | 0.228 |
| PROTEIN | Total protein | 0.104 | 1.386 | 0.229 |
| K. | Potassium | 0.109 | 1.455 | 0.229 |
| w_age | Maternal age at inclusion | 0.112 | 1.498 | 0.239 |
| FGF.21 | Fibroblast growth factor 21 | 0.104 | 1.385 | 0.242 |
| vitamin.A | Vitamin A | 0.105 | 1.397 | 0.258 |
| TrpBetaine | Tryptophan betaine | 0.104 | 1.389 | 0.271 |
| X1.Met.His | Methylhistidine | 0.102 | 1.367 | 0.275 |
| DG.14.0_18.1. | Diglyceride (14:0/18:1) | 0.097 | 1.300 | 0.290 |
| PLP | Pyridoxal-5-phosphate | 0.092 | 1.234 | 0.315 |
| LNnT | Lacto-*N*-neotetraose | 0.093 | 1.247 | 0.315 |
| Sia | HMO bound sialic acid | 0.086 | 1.149 | 0.318 |
| FLNH | Fucosyllacto-*N*-hexaose | 0.090 | 1.201 | 0.324 |
| Collinsella | Milk genus Collinsella | 0.093 | 1.243 | 0.331 |
| DSLNH | Disialyllacto-*N*-hexaose | 0.089 | 1.192 | 0.336 |
| season_lean | Infant born in lean season (yes/no) | 0.079 | 1.049 | 0.361 |
| TG.18.3_32.0. | Triglyceride (18:3/32:0) | 0.082 | 1.089 | 0.363 |
| DFLNT | Difucosyllacto-*N*-tetraose | 0.082 | 1.095 | 0.368 |
| PC.ae.C40.4 | Phosphatidylcholine acyl-alkyl C40:4 | 0.082 | 1.098 | 0.382 |
| Hex2Cer.d18.1.24.0. | Hexosylceramide (d18:1/24:0) | 0.076 | 1.011 | 0.383 |
| NAM | Nicotinamide | 0.075 | 1.003 | 0.399 |
| TPP | Thiamine pyrophosphate | 0.077 | 1.022 | 0.406 |
| GWG | Gestational weight gain | 0.079 | 1.055 | 0.411 |
| Code | Maternal balanced energy–protein supplementation (yes/no) | 0.075 | 1.001 | 0.423 |
| LH | Luteinizing hormone | 0.073 | 0.970 | 0.425 |
| Cer.d18.1.16.0. | Ceramide (d18:1/16:0) | 0.071 | 0.954 | 0.434 |
| Veillonella_A | Milk genus *Veillonella* A | 0.065 | 0.866 | 0.472 |
| m_weightincl | Maternal weight at inclusion | 0.067 | 0.901 | 0.473 |
| parity | Number of pregnancies | 0.064 | 0.849 | 0.475 |
| LNT | Lacto-*N*-tetrose | 0.068 | 0.905 | 0.480 |
| asset1comp_10 | Maternal wealth index (1 to 10) | 0.065 | 0.873 | 0.482 |
| LNFP.II | Lacto-*N*-fucopentaose II | 0.060 | 0.806 | 0.500 |
| g.tocopherol | Gamma-tocopherol (Vitamin E) | 0.063 | 0.844 | 0.503 |
| TRP | Tryptophan | 0.060 | 0.804 | 0.511 |
| Zn | Zinc | 0.058 | 0.778 | 0.530 |
| m_heightincl | Maternal height at inclusion | 0.055 | 0.740 | 0.542 |
| Insulin | Insulin | 0.051 | 0.681 | 0.591 |
| LNH | Lacto-*N*-hexaose | 0.047 | 0.630 | 0.631 |
| PL | Pyridoxal | 0.045 | 0.597 | 0.660 |
| m_muacincl | Maternal mid-upper arm circumference at inclusion | 0.043 | 0.578 | 0.661 |
| Leptin | Leptin | 0.041 | 0.553 | 0.678 |
| LNFP.I | Lacto-*N*-fucopentaose I | 0.042 | 0.566 | 0.684 |
| DFLac | Difucosyllactose | 0.038 | 0.510 | 0.739 |
| m_bmiincl | Maternal body mass index at inclusion | 0.035 | 0.470 | 0.743 |
| Parolsenella | Milk genus *Parolsenella* | 0.033 | 0.434 | 0.772 |
| PN | Pyridoxine | 0.022 | 0.290 | 0.912 |

**Table S6. Complete list of all estimated paths from the final structural equation model, related to Figure 7**

| Dependent (outcome) variable | op | Independent (predictor) variable | Estimate | Standardized estimate | p-value |
|---|---|---|---|---|---|
| Infant_Gut_12m | ~ | Milk_Metabolite_1421d | 0.140 | 0.140 | 0.277 |
| Infant_Gut_12m | ~ | Milk_Nutrient_1421d | -0.105 | -0.105 | 0.370 |
| Infant_Gut_12m | ~ | Milk_Microbiome_12m | -0.557 | -0.557 | 3.33e-15 |
| Infant_Gut_12m | ~ | Milk_Metabolite_12m | -0.099 | -0.099 | 0.168 |
| Infant_Gut_12m | ~ | Mother_Gut_12m | 0.082 | 0.082 | 0.195 |
| Infant_Gut_12m | ~ | Mother_Gut_Tri3 | 0.620 | 0.620 | 0 |
| Infant_Gut_56m | ~ | Milk_Microbiome_1421d | -0.345 | -0.328 | 3.456 |
| Infant_Gut_56m | ~ | Milk_Metabolite_34m | -0.739 | -0.703 | 1.51e-11 |
| Infant_Gut_56m | ~ | Milk_Nutrient_34m | -0.255 | -0.242 | 0.015 |
| Infant_Gut_56m | ~ | Mother_Gut_56m | -0.594 | -0.565 | 0 |
| Milk_Metabolite_34m | ~ | Infant_Gut_12m | 0.233 | 0.233 | 6.48e-09 |
| Milk_Metabolite_34m | ~ | Milk_Nutrient_34m | -0.748 | -0.748 | 0 |
| Infant_Gut_56m | ~ | Milk_Metabolite_1421d | 0.750 | 0.714 | 0 |
| Milk_Metabolite_34m | ~ | Milk_Microbiome_1421d | -0.237 | -0.237 | 3.986e-07 |
| Infant_Gut_56m | ~ | Milk_Metabolite_12m | 0.248 | 0.236 | 0.0006 |
| Infant_Gut_56m | ~ | Mother_Gut_12m | -0.161 | -0.154 | 0.002 |
| Milk_Metabolite_34m | ~ | Milk_Nutrient_1421d | 0.233 | 0.234 | 1.42e-07 |
| Milk_Metabolite_34m | ~ | Milk_Metabolite_12m | 0.235 | 0.235 | 0.0002 |
| Infant_Gut_56m | ~ | Infant_Gut_12m | -0.432 | -0.412 | 2.2e-09 |
| Infant_Gut_56m | ~ | Mother_Gut_Tri3 | 0.420 | 0.399 | 1.65e-10 |
| Infant_Gut_56m | ~ | Milk_Microbiome_12m | -0.333 | -0.317 | 1.4e-07 |
| Infant_Gut_56m | ~ | Milk_Nutrient_1421d | -0.263 | -0.250 | 0.002 |
| Infant_Gut_12m | ~ | Milk_Microbiome_1421d | -0.129 | -0.129 | 0.078 |
| Infant_Gut_12m | ~~ | Infant_Gut_12m | 0.431 | 0.434 | 0 |
| Infant_Gut_56m | ~~ | Infant_Gut_56m | 0.215 | 0.196 | 1.78e-15 |
| Milk_Metabolite_34m | ~~ | Milk_Metabolite_34m | 0.193 | 0.195 | 2.22e-16 |

~ indicates a regression path, meaning “was predicted by”.

~~ indicates a covariance (or residual variance), meaning “residual variance of”

Time point abbreviation: 1421d, 14–21 days; 12m, 1–2 months; 34m, 3–4 months; 56m, 5–6 months
